# Mapping the Health Burden of Neighbourhood Deprivation: Neurobiological Evidence Across the Life Span

**DOI:** 10.64898/2026.08.29.26361714

**Authors:** Amir Ebneabbasi, Varun Warrier, Marcella Montagnese, Rafael Romero Garcia, Richard A.I. Bethlehem, Timothy Rittman

## Abstract

Neighbourhood deprivation is one of the few potential policy-modifiable risk factors for psychiatric and neurological disorders, but the neurobiological pathways underlying these associations remain unclear. We investigated these relationships across three cohorts spanning the life span: the Healthy Brain and Child Development (HBCD) Study (n = 84, aged 0–4 weeks postnatal), the Adolescent Brain Cognitive Development (ABCD) Study (n = 4,792, aged 9–10 years), and the UK Biobank (UKB; ∼ 500,000 adults, aged 44–87 years). Neighbourhood deprivation was associated with elevated disease risk, and individual lifestyle factors accounted for only a small fraction of this burden, indicating that the much larger residual effect reflects broader contextual characteristics of deprived environments rather than individual behaviours alone. Across all cohorts, greater deprivation consistently predicted lower cortical and subcortical brain volume, with effects detectable in early development and substantially stronger in adulthood. Across disorders, regional brain volume emerged as a consistent neuroanatomical mediator linking neighbourhood deprivation to neuropsychiatric disease. We further showed that deprivation preferentially affects brain regions intrinsically vulnerable to neuropsychiatric disorders. Spatial decoding analyses implicated dopaminergic and serotonergic neurotransmitter systems together with specific excitatory and inhibitory neuronal classes. Importantly, both the deprivation–disease associations and their neuroanatomical mediation patterns replicated across independent populations. Our study delivers a translational framework linking neighbourhood deprivation to brain health which could inform public health policies and preventive interventions.

## Introduction

Measures of neighbourhood deprivation have emerged as powerful predictors of health [1–3] and mortality [4]. Composite indices such as the Area Deprivation Index (ADI) in the United States and the Index of Multiple Deprivation (IMD) in the United Kingdom complement traditional physical-environment measures by capturing the socioeconomic aspects of living environments. Governments use these indices to identify priority areas across sectors, including healthcare, education, housing, and social services [5, 6]. Quantifying the cumulative effects represented by these indices is therefore essential not only for understanding disease mechanisms but also for evaluating and informing public policy decisions.

Observational studies have demonstrated that neighbourhood deprivation, measured using the ADI or IMD, is associated with an increased risk of individual psychiatric [7, 8] or neurological disorders [9, 10]. Although lifestyle factors are proposed as potential mediators of socioeconomic health inequalities [11, 12], the extent to which deprivation-associated disease risk reflects individual lifestyle behaviours, rather than broader neighbourhood-level influences, remains largely unknown. This distinction is important because it informs whether policies should prioritise individual behaviour change or broader neighbourhood-level interventions.

Neuroimaging studies have shown that neighbourhood deprivation is associated with regional differences in brain volume [13–16], but it remains unknown whether brain structure mediates the relationship between deprivation and specific disorders. If so, which developmental periods and disorder categories are most vulnerable to the neurobiological effects of socioeconomic disadvantage remains an important question about when and where preventive interventions may be most effective.

Beyond identifying deprivation-related brain alterations, it remains unclear whether socioeconomic disadvantage acts through neurobiological systems already implicated in disease or through distinct anatomical pathways. Likewise, the broader neurobiological context of deprivation-related brain alterations remains poorly understood [17, 18], including whether they align with established principles of brain organisation, such as large-scale functional networks, neurotransmitter systems, and cellular architecture. Characterising these relationships could identify potential targets for future interventions.

To address these gaps, we conducted a comprehensive life span investigation of the relationships between neighbourhood deprivation, brain structure, and neuropsychiatric disease using three population-based cohorts spanning development to older adulthood: the Healthy Brain and Child Development (HBCD) Study (n = 84, aged 0–4 weeks postnatal) [19], the Adolescent Brain Cognitive Development (ABCD) Study (n = 4,792 children aged 9–10 years) [20], and the UK Biobank (UKB; ∼500,000 adults aged 44-87 years) [21]. We quantified the effects of deprivation across a broad range of psychiatric and neurological disorders, assessed the contribution of individual lifestyle factors, mapped deprivation-related brain volume differences across development, and tested whether these neuroanatomical alterations mediate disease risk. We then examined whether deprivation disproportionately affects regions intrinsically vulnerable to neuropsychiatric disorders and contextualised these patterns using biologically informative brain maps. Finally, we evaluated the robustness and generalisability of all findings through replication and targeted sensitivity analyses addressing temporal ordering and potential confounding, including population stratification and passive gene–environment correlation.

## Results

### Deprivation is associated with increased risk of neuropsychiatric disorders across the lifespan

The effect of neighbourhood deprivation on the occurrence of neuropsychiatric disorders was examined using a binomial generalised linear model (GLM). All regression models were bootstrapped with 5,000 resamples to obtain robust parameter estimates and 95% confidence intervals. Analyses were restricted to individuals of European ancestry and adjusted for genetic, demographic, and neuroimaging covariates (see Methods). Only the ABCD and UKB cohorts were included for this analysis, since HBCD participants have not yet reached an age at which reliable diagnostic ascertainment is possible.

We selected composite measures of neighbourhood deprivation that are broadly comparable across cohorts. Specifically, the Area Deprivation Index (ADI) in ABCD and the Index of Multiple Deprivation (IMD) in UKB-England were used as the primary predictors in regression analyses. Because socioeconomic disadvantage rarely occurs along a single dimension, composite deprivation indices integrate information across multiple socioeconomic domains and may therefore capture the broader context of cumulative disadvantage more comprehensively than individual indicators [22]. To better characterise deprivation indices in our work, we examined their correlations with the individual socioeconomic indicators from which they were derived (Extended Data Figure 1).

Diagnostic outcomes were restricted to those with at least 50 cases, resulting in 24 K-SADS-PL DSM-5 psychiatric diagnoses in ABCD (N controls = 416) and 109 ICD-10 diagnoses in UKB-England (N controls = 225,877), comprising 32 mental and behavioural disorders (ICD-F) and 77 diseases of the nervous system (ICD-G). An overview of the study design is provided in Figure 1. The total effects of neighbourhood deprivation on disease outcomes are presented in Figure 2 and Supplementary Tables 1 (ABCD) and 2 (UKB).

**Figure 1.**
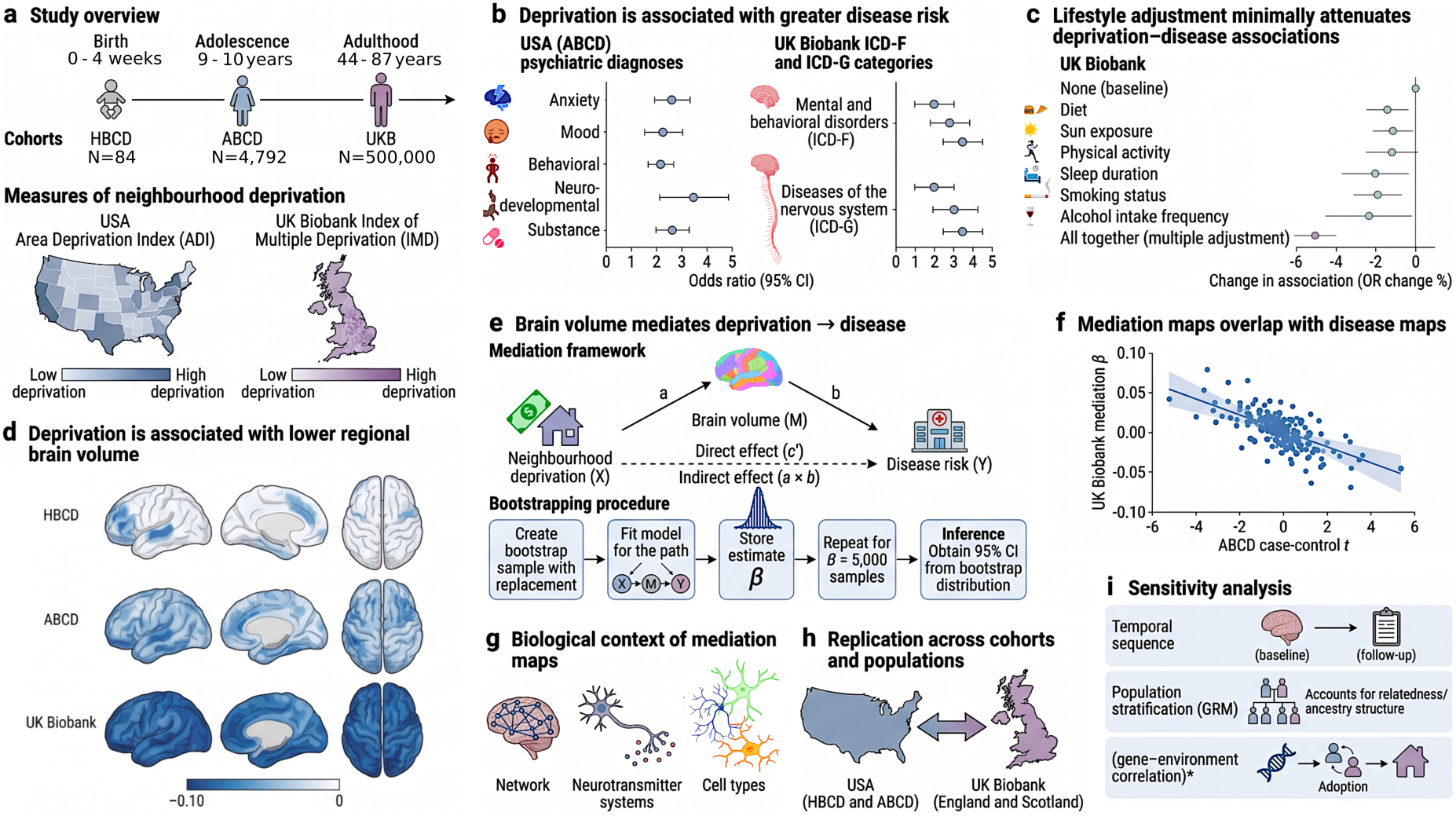
Study overview and analytical framework. a, Neighbourhood deprivation was examined across the life span using HBCD (0–4 weeks; n = 84), ABCD (9–10 years; n = 4,792), and the UK Biobank (44-87 years; ∼500,000), using the US Area Deprivation Index (ADI) and UK Index of Multiple Deprivation (IMD). **b,** Associations between deprivation and psychiatric and neurological disease risk were assessed across diagnostic categories. **c,** Lifestyle adjustment tested the contribution of diet, sun exposure, physical activity, sleep, smoking, and alcohol use to deprivation–disease associations. **d,** Regional brain-volume associations with deprivation were mapped across infancy, childhood, and adulthood. **e,** Mediation analyses tested whether deprivation-related differences in brain volume mediate disease risk. **f,** Mediation maps were compared with neuroanatomical patterns associated with neuropsychiatric disorders. **g,** Their biological context was characterised using functional networks, neurotransmitter systems, and cellular architecture. **h,** Findings were evaluated for replication and generalisability across cohorts and populations. **i,** Sensitivity analyses addressed temporal ordering, population stratification, and passive gene–environment correlation. The figure was created by BioRender.com.

**Figure 2.**
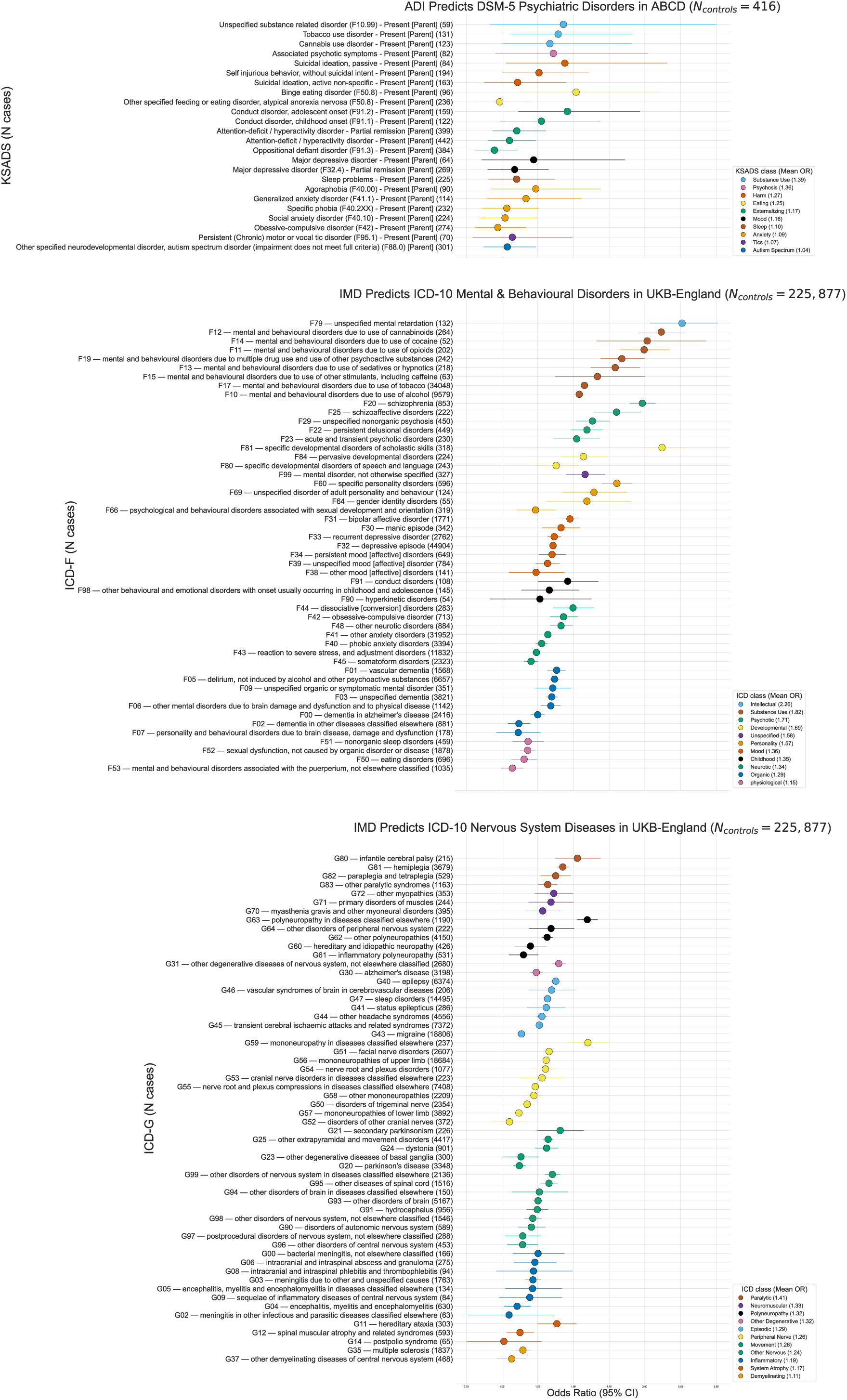
Total effects of neighbourhood deprivation on psychiatric and neurologic diseases in ABCD and UKB-England. All models were bootstrapped (5,000 resamples) to obtain robust parameter estimates and 95% confidence intervals. Analyses were restricted to individuals of European ancestry and adjusted for genetic, demographic, and neuroimaging covariates (Methods). Neighbourhood deprivation was indexed using the Area Deprivation Index (ADI) in ABCD and the Index of Multiple Deprivation (IMD) in UKB-England, both of which are composite measures capturing multiple dimensions of socioeconomic disadvantage. Diagnostic categories were limited to those with ≥50 cases, yielding 24 K—SADS-PL DSM-5 diagnoses in ABCD (N controls = 416) and 109 ICD-10 diagnoses in UKB-England (N controls = 225,877), comprising 32 ICD-F and 77 ICD-G disorders. Total effects of neighbourhood deprivation on each diagnostic outcome are shown.

In the ABCD cohort, greater deprivation (higher ADI score) was associated with increased odds across several psychiatric domains. The strongest associations were observed for substance use disorders (mean OR = 1.39, range = 1.34–1.43), followed by psychotic disorders (OR = 1.36), harm-related symptoms (mean OR = 1.27, range = 1.11–1.44), eating disorders (mean OR = 1.25, range = 0.98–1.52), and externalising disorders (mean OR = 1.17, range = 0.95–1.46). See Figure 2 for the complete set of associations.

In the UKB-England cohort, greater deprivation (higher IMD) was associated with increased odds across all major ICD-10 mental disorder categories. The strongest associations were observed for mental retardation (intellectual disability) (OR = 2.26), followed by mental and behavioural disorders due to psychoactive substance use (mean OR = 1.82, range = 1.54–2.12), schizophrenia, schizotypal and delusional disorders (mean OR = 1.71, range = 1.52–1.98), disorders of psychological development (mean OR = 1.69, range = 1.38–2.12), and disorders of adult personality and behaviour (mean OR = 1.57, range = 1.24–1.81). Figure 2 provides a complete summary of the observed associations.

Among ICD-10 neurological disorders, greater deprivation (higher IMD) was likewise associated with increased odds across multiple disease categories. The strongest associations were observed for cerebral palsy and other paralytic syndromes (mean OR = 1.41, range = 1.32–1.53), followed by diseases of the myoneural junction and muscle (mean OR = 1.33, range = 1.29–1.36), polyneuropathies and other disorders of the peripheral nervous system (mean OR = 1.32, range = 1.15–1.60), other degenerative diseases of the nervous system (mean OR = 1.32, range = 1.24– 1.40), and episodic and paroxysmal disorders (mean OR = 1.29, range = 1.14–1.38). The complete set of associations can be found in Figure 2.

### Deprivation–disease associations are largely preserved after lifestyle adjustment

A key question is whether the association between neighbourhood deprivation and disease is attributable to differences in individual lifestyle behaviours or remains evident after accounting for these factors. To examine this, we repeated the deprivation–disease analyses with additional adjustment for lifestyle variables while retaining the baseline covariates. This analysis was intended to assess the extent to which the estimated association between deprivation and disease was attenuated after adjustment for measured lifestyle behaviours. The analysis was restricted to the UKB-England cohort because participants in the ABCD study were aged 9–10 years and had not yet developed the broad range of adult lifestyle behaviours examined here, including alcohol intake, smoking status, diet, sun exposure, sleep duration, and physical activity. Models were compared by the percentage attenuation of the IMD regression coefficient (see Methods).

Simultaneous adjustment for all lifestyle factors resulted in only modest attenuation of the association between IMD and disease, reducing the estimated deprivation effect by 6.34% for psychiatric disorders and 5.16% for neurological disorders. Among the individual lifestyle variables, adjustment for alcohol intake yielded the largest attenuation (mean reduction, 3.19% for psychiatric disorders and 3.04% for neurological disorders), followed by smoking status (2.17% and 1.43%, respectively). Importantly, the majority of associations that were significant in the baseline analyses remained statistically significant after lifestyle adjustment (Figure 3 and Supplementary Table 3). These findings suggest that deprivation-related disease risk is largely shaped by factors beyond the measured individual lifestyle behaviours, highlighting the potential importance of broader socioeconomic and psychosocial mechanisms.

**Figure 3.**
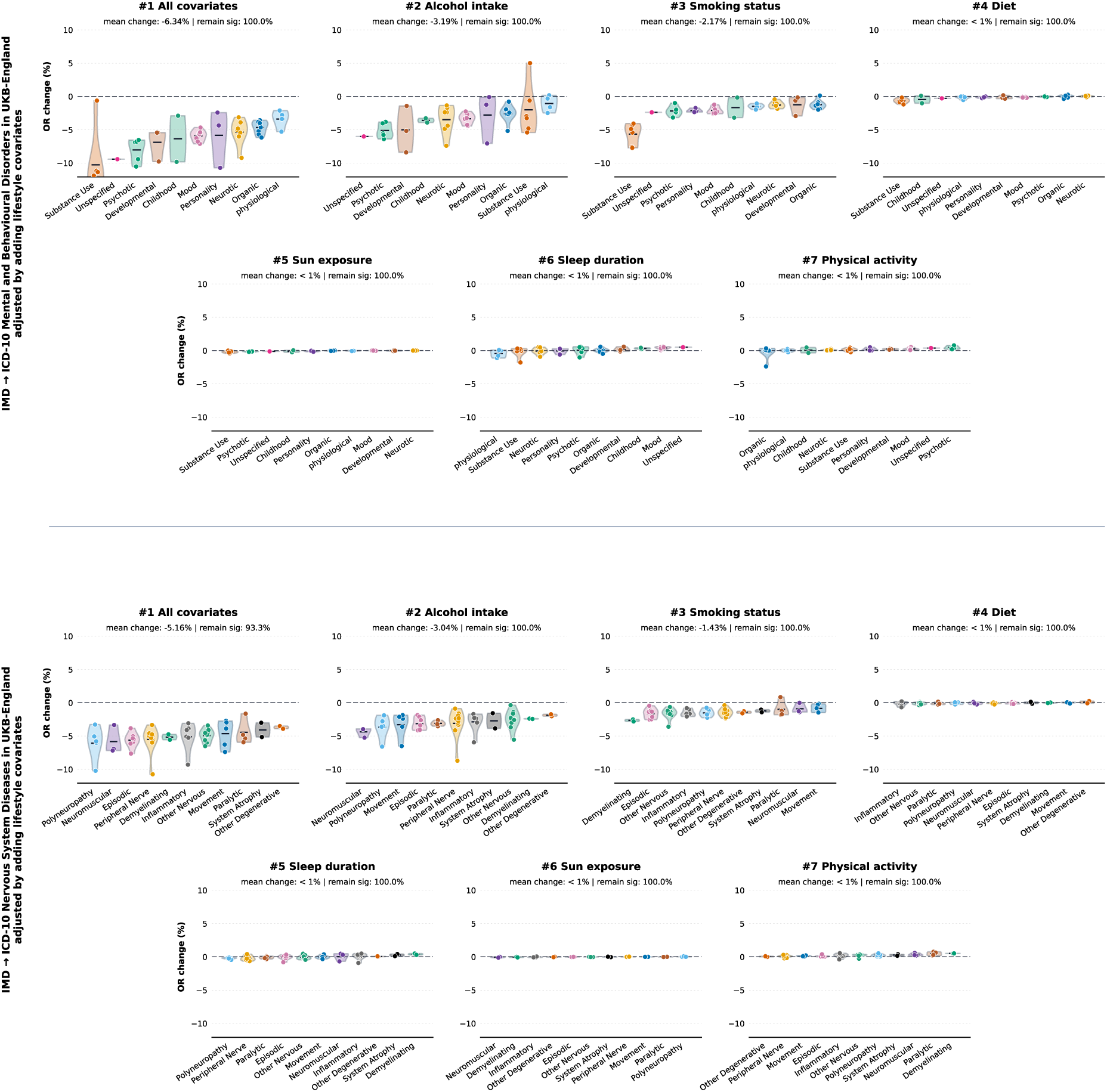
Lifestyle-adjusted deprivation–disease association analysis in UKB-England. Percentage change in the Odds ratio for the association between Index of Multiple Deprivation (IMD) and individual ICD-10 psychiatric disorders (top) and neurological disorders (bottom) after additional adjustment for lifestyle factors, relative to models containing the baseline covariates. Points represent individual disorders, and distributions summarise variation across categories. Negative values indicate attenuation of the IMD–disease association following lifestyle adjustment, whereas values close to zero indicate little change relative to the baseline model. The text above each panel reports the mean percentage change in the IMD coefficient and the proportion of associations that remained statistically significant after additional adjustment.

### Deprivation is associated with reduced brain volume across the lifespan

We investigated the association between neighbourhood deprivation and regional brain volumes across three cohorts using ordinary least squares (OLS) regression. Neighbourhood deprivation was quantified using the ADI in the HBCD (n = 84) and ABCD (n = 4,792) cohorts, and the IMD in the UKB-England (n = 63,197) cohort. We used bilaterally averaged measures from 34 cortical and cohort-specific subcortical regions (6 in HBCD; 9 in ABCD and UKB-England). Because T2-weighted MRI provides superior tissue contrast in the neonatal brain, regional measures in HBCD were derived from T2-weighted images processed with fMRIPrep Lifespan [23] (Methods). In contrast, regional measures in ABCD and UKB-England were derived from T1-weighted images processed with FreeSurfer [24]. Analyses were restricted to participants of European ancestry and adjusted for genetic, demographic, and neuroimaging covariates (Methods).

The direction of association was largely consistent across all cohorts, indicating that greater neighbourhood deprivation was associated with reduced brain volume throughout the cortex and subcortex, with the strongest and most widespread effects observed in the larger and older cohorts (Figure 4 and Supplementary Table 4). In the HBCD cohort, no regional brain volumes were significantly associated with ADI after adjusting for the Benjamini–Hochberg false discovery rate (FDR). This may reflect the limited statistical power of the relatively small sample and/or the early developmental stage of the participants, during which the cumulative effects of socioeconomic deprivation on brain structure may not yet be fully detectable. In contrast, in the ABCD cohort, higher ADI was significantly associated with lower volumes across all regions (P_FDR_ < 0.05), except the isthmus cingulate, pars opercularis, transverse temporal, pars triangularis, rostral anterior cingulate, and insular cortices. Similarly, in the UKB, higher IMD was robustly associated with smaller volumes across all cortical and subcortical regions examined, with consistent negative standardised regression coefficients and highly significant FDR-corrected p-values.

**Figure 4.**
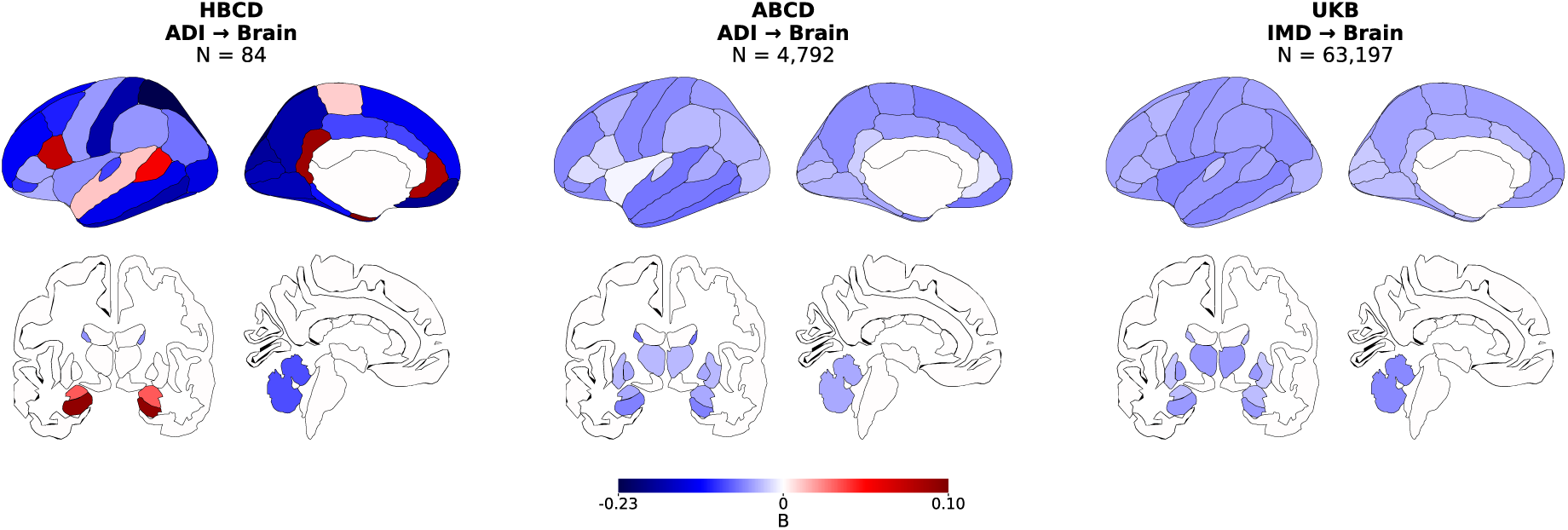
Effects of neighbourhood deprivation on subcortical and cortical volumes across HBCD, ABCD and UKB. Brain maps show standardised regression coefficients (β) from ordinary least squares (OLS) models examining associations between neighbourhood deprivation and regional cortical and subcortical brain volumes in HBCD (ADI; n = 84), ABCD (ADI; n = 4,792), and UKB-England (IMD; n = 63,197). Regional measures comprised 34 cortical regions and cohort-specific subcortical regions (6 in HBCD and 9 in ABCD and UKB-England). HBCD measures were derived from T2-weighted MRI, whereas ABCD and UKB-England measures were derived from T1-weighted MRI (Methods). Models were restricted to participants of European ancestry and adjusted for genetic, demographic, and neuroimaging covariates. Negative β values (blue) indicate that greater neighbourhood deprivation was associated with smaller regional brain volumes.

### Brain volume mediates the effect of deprivation on disease risk

We next investigated whether regional brain volumes mediate the associations between neighbourhood deprivation and disease outcomes. Only ABCD and UKB were included, as reliable diagnosis is not yet possible in HBCD. Diagnostic outcomes were restricted to those with at least 50 cases, resulting in 24 KSADS-5 psychiatric diagnoses in ABCD (N controls = 416) and 48 ICD-10 diagnoses in UKB-England (N controls = 39,789), comprising 25 mental and behavioural disorders (ICD-F) and 23 diseases of the nervous system (ICD-G). The indirect effect was calculated as the product of path *a*, estimated using OLS linear regression (deprivation → brain volume), and path *b*, estimated using binomial GLM (brain volume → disease outcome, adjusted for deprivation) (Methods).

The findings were remarkably consistent across both the ABCD and UKB-England cohorts. Positive indirect effects were observed for the vast majority of diseases, reflecting the combination of two negative associations: greater deprivation was associated with lower regional brain volume, which in turn was associated with higher disease risk (Figure 5). Results are also provided in Supplementary Tables 5 (ABCD) and 6 (UKB).

**Figure 5.**
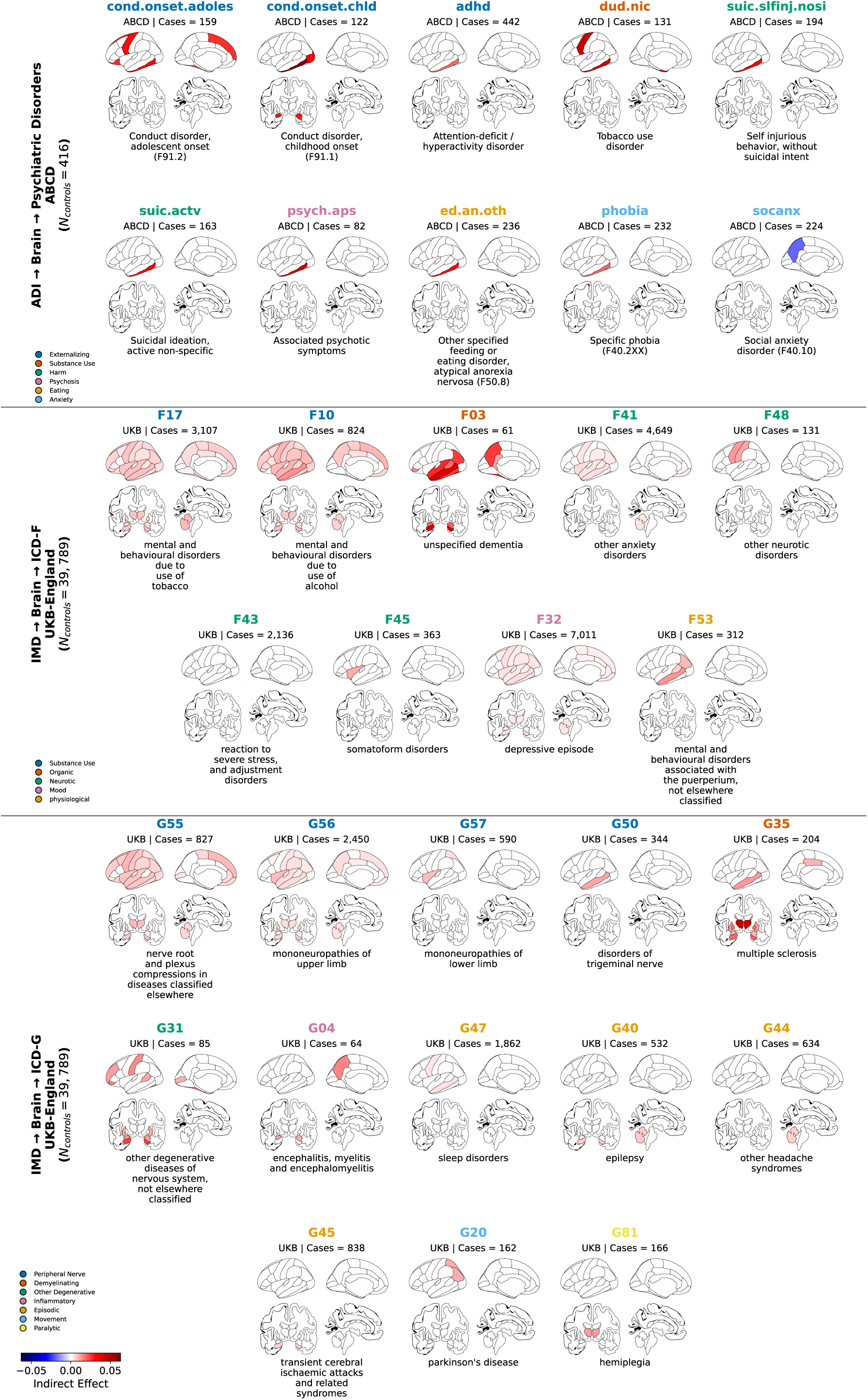
Regional brain volumes mediate associations between neighbourhood deprivation and psychiatric and neurological disorders. Brain maps show regional indirect effects of neighbourhood deprivation on psychiatric disorders in ABCD (top), and mental and behavioural disorders (ICD-10 Chapter F; middle) and diseases of the nervous system (ICD-10 Chapter G; bottom) in UKB-England. Mediation was quantified as the product of path a (deprivation → regional brain volume), estimated using OLS regression, and path b (regional brain volume → disease outcome, conditional on deprivation), estimated using binomial GLM. Analyses included diagnostic outcomes with at least 50 cases. In ABCD, disorders with at least one nominally significant regional indirect effect (P < 0.05) are displayed; none survived FDR correction. In UKB-England, only disorders with at least one regional indirect effect surviving FDR correction (P_FDR_ < 0.05) are shown. Coloured brain regions indicate significant mediating regions, with colour intensity representing the magnitude and direction of the indirect effect (blue, negative; red, positive); uncoloured regions did not meet the corresponding significance threshold. Disorder-label colours indicate diagnostic domains, as defined in the accompanying legends. Positive indirect effects predominated across both cohorts, consistent with greater neighbourhood deprivation being associated with smaller regional brain volumes, which in turn were associated with greater disease risk.

In the ABCD dataset, disorders were plotted in Figure 5 for which at least one brain region showed a nominally significant indirect effect (P < 0.05), as no associations survived FDR correction. Significant mediation of brain structure on the effect of deprivation was identified for 10 psychiatric phenotypes spanning six domains. The Externalising domain, comprising conduct disorder (childhood- and adolescent-onset) and attention-deficit/hyperactivity disorder, exhibited the largest indirect effects (β = 0.018–0.063), mediated by the inferior temporal, precentral, superior frontal, lateral orbitofrontal, lateral occipital, and fusiform cortices, as well as the amygdala. The Substance Use domain, represented by tobacco use disorder, showed indirect effects ranging from β = 0.025 to 0.044 and implicated the inferior temporal, precentral, entorhinal, and temporal pole cortices. Overall, the inferior temporal cortex was the most prominent mediator, significantly transmitting deprivation-related risk to nine disorders, including those in the Harm domain, represented by suicidal ideation and non-suicidal self-injury (β = 0.032–0.033), associated psychotic symptoms (β = 0.042), and atypical anorexia nervosa (β = 0.030). In contrast, the effect of deprivation on the Anxiety domain, comprising social anxiety disorder and specific phobia (β = −0.019 to 0.019), was mediated by the inferior temporal cortex and precuneus and was the only domain to exhibit both positive and negative indirect effects.

Within ICD-10 Chapter F in the UKB-England, the mediatory role of brain structure varied in spatial extent and magnitude. Diseases were plotted in Figure 5 that exhibited at least one brain region with a significant indirect effect after correction for multiple comparisons (P_FDR_ < 0.05). Significant mediation by brain structure in the effect of deprivation was observed for nine disorders across five domains. The Substance Use domain, comprising alcohol- and tobacco-related disorders, showed indirect effects of brain structure ranging from β = 0.001 to 0.013 across lateral and ventral temporal cortices with limbic and subcortical structures. The Organic domain, including different types of dementia and cognitive impairment secondary to brain disease, exhibited the largest and most spatially extensive indirect effects (β = 0.010–0.040). Widespread cortical areas were implicated, including temporal and inferior parietal cortices, as well as extensive subcortical and cerebellar regions. The Neurotic domain, including anxiety, stress-related, somatoform, and other neurotic disorders (β = −0.001 to 0.013), demonstrated more localised mediation involving temporal cortical regions, as well as the hippocampus and amygdala. The Mood domain, represented by depressive episode (β = 0.001–0.004), showed comparatively modest mediation that was largely restricted to temporal and limbic regions, whereas the Physiological domain, represented by puerperal mental and behavioural disorders (β = 0.006–0.013), exhibited focal mediation mainly within the ventral temporal cortex.

Within ICD-10 Chapter G, 13 neurological disorders exhibited at least one significant indirect effect following FDR correction (P _FDR_ < 0.05). These disorders represented seven ICD-G domains. The Peripheral Nerve class, including mononeuropathies and nerve root disorders (β = 0.001– 0.011), demonstrated mediation of the effect of deprivation by volumes of temporal and parietal cortices, together with selected subcortical structures. The Other Degenerative domain (β = 0.010–0.024) and the Demyelinating domain, represented by multiple sclerosis (β = 0.009– 0.045), exhibited widespread involvement of subcortical, cerebellar, and temporal regions. The Inflammatory domain (β = 0.011–0.015) displayed a similar but more restricted cortico-subcortical pattern. The Episodic domain, including epilepsy, headache syndromes, sleep disorders, and transient cerebral ischaemic attacks (β = −0.002 to 0.007), showed comparatively focal mediation involving limbic and temporal regions. The Movement domain, represented by Parkinson’s disease (β = 0.010), and the Paralytic domain, represented by hemiplegia (β = 0.012), were dominated by subcortical regions with relatively limited cortical involvement.

### Deprivation preferentially targets disease-vulnerable brain regions

The next question we sought to address was whether neighbourhood deprivation preferentially affects brain regions that are intrinsically vulnerable to disease. We compared the mediation maps derived from UKB–England with independent case–control maps. The cross-cohort analyses included five disorders in ABCD, as equivalent phenotypic matches were unavailable for all disorders, and four diseases in UKB-Scotland, as not all diseases had sufficient case numbers in the Scottish cohort. The procedure used to generate the case–control maps is described in the Methods, and the resulting maps are provided in Supplementary Tables 7 and 8 for ABCD and UKB–Scotland, respectively. Spatial co-locations involving cortical maps were assessed using 1,000 Spin permutations [25], and both cortical and subcortical results were corrected for multiple comparisons (Figure 6).

**Figure 6.**
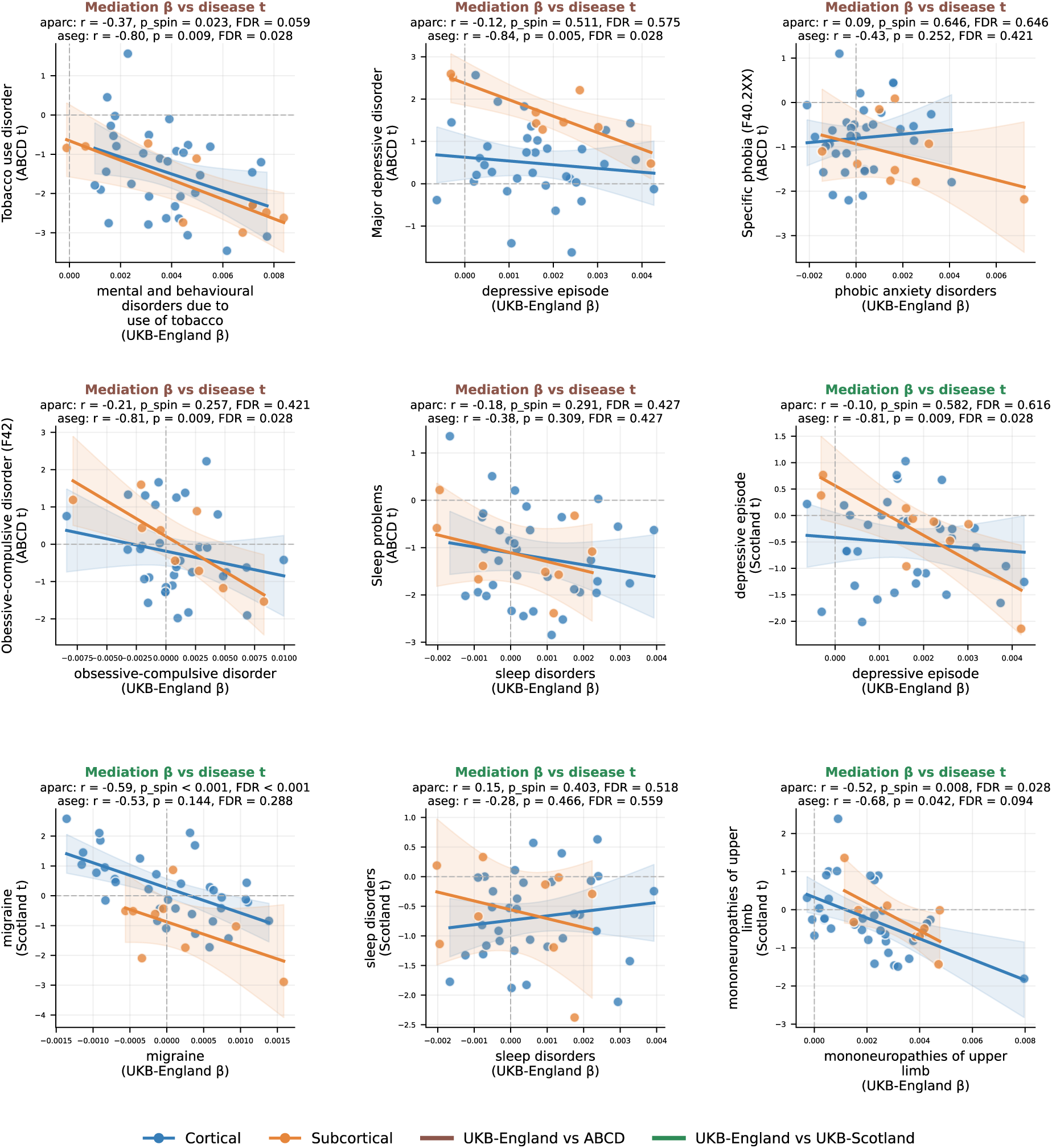
Spatial correspondence between disease-related t-maps and deprivation-related mediation β-maps, demonstrating that anatomical changes implicated in deprivation overlap with brain regions susceptible to neuropsychiatric disorders. Scatterplots show spatial correlations between regional mediation effects (β) derived from the UKB-England deprivation–brain–disease analyses and independent case–control effect-size maps (t-statistics) from ABCD or UKB-Scotland. Each point represents a cortical (blue) or subcortical (orange) region, with fitted regression lines and 95% confidence intervals shown separately for the two anatomical parcellations. Negative correlations indicate that regions showing stronger positive mediation effects of neighbourhood deprivation tended to exhibit greater disease-related reductions in brain volume. Cortical spatial correspondence was assessed using 1,000 Spin permutations, whereas subcortical associations were evaluated using conventional correlations; results were corrected for multiple comparisons using the Benjamini–Hochberg false discovery rate (FDR).

Across nine conditions and two anatomical parcellations, 16 of 18 spatial correlations were negative, indicating that brain regions showing stronger mediation of neighbourhood deprivation tended to exhibit greater disease-related volumetric loss. In the UKB-England versus ABCD comparisons, tobacco use showed significant spatial co-localisation in both the cortex (*r* = −0.37, P_FDR-Spin_ = 0.059) and subcortex (*r* = −0.80, P_FDR_ = 0.028). Significant subcortical associations were also observed for depressive episodes (*r* = −0.84, P_FDR_ = 0.028) and obsessive–compulsive disorder (*r* = −0.81, P_FDR_ = 0.028). The remaining UKB-England versus ABCD comparisons did not show significant spatial co-localisation (Figure 6).

In the UKB-England versus UKB-Scotland comparisons, migraine exhibited a strong cortical association (*r* = −0.59, P_FDR-Spin_ < 0.001), while depressive episode showed a subcortical association (*r* = −0.81, P_FDR_ = 0.028). Mononeuropathies of the upper limb demonstrated significant cortical co-localisation (*r* = −0.52, P_FDR-Spin_ = 0.028), whereas the corresponding subcortical association was nominally significant (*r* = −0.68, P= 0.042). The remaining disorders did not show significant spatial co-localisation. Together, these findings indicate that neighbourhood deprivation disproportionately affects brain systems that are susceptible to neuropsychiatric disorders (Figure 6).

### Deprivation-related brain changes map onto established neurobiological architecture

To place deprivation-related brain changes within a broader neurobiological context, we performed spatial decoding analyses using (1) the Yeo large-scale functional networks [26]; (2) PET-derived maps of 19 neurotransmitter receptors, transporters, and binding sites spanning nine neurotransmitter systems [27]; and (3) cell-type abundance maps derived from the Allen Human Brain Atlas [28] with markers obtained from single-nucleus RNA sequencing (snRNA-seq) data, encompassing 24 cell classes (nine inhibitory neuronal, nine excitatory neuronal, and six non-neuronal classes) [29]. Spatial co-localisations involving cortical maps were assessed using 1,000 Spin permutations, and all analyses were corrected for multiple comparisons (Figure 7 and Supplementary Table 9).

**Figure 7.**
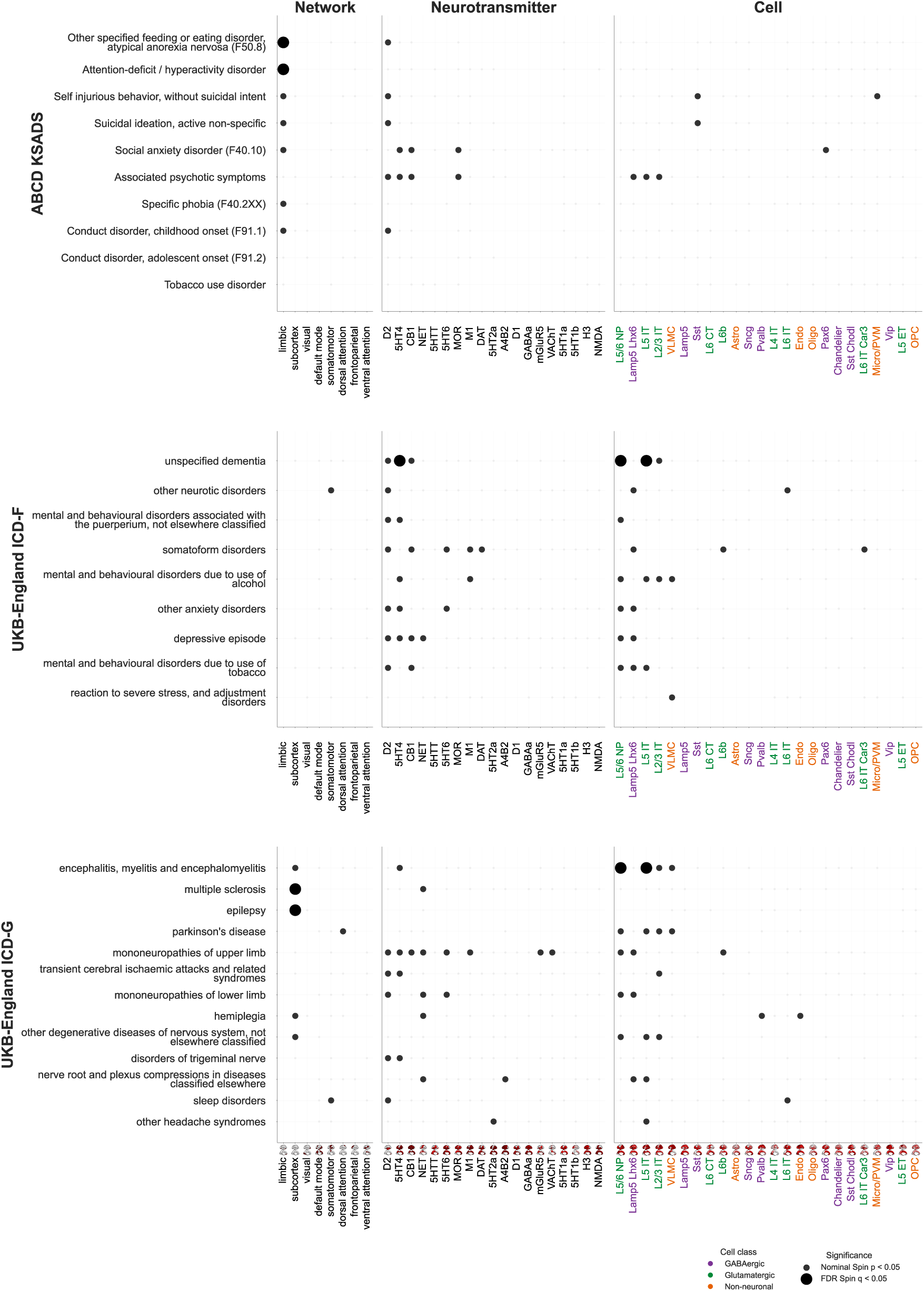
Neurobiological decoding of brain regions mediating associations between neighbourhood deprivation and psychiatric and neurological disorders. Spatial decoding analyses characterise the neurobiological organisation of deprivation-related mediation maps for psychiatric disorders in ABCD (top), and mental and behavioural disorders (ICD-10 Chapter F; middle) and diseases of the nervous system (ICD-10 Chapter G; bottom) in UKB-England. Mediation maps were spatially compared with three complementary neurobiological reference systems: Yeo large-scale functional networks (Network), PET-derived maps of 19 neurotransmitter receptors, transporters, and binding sites spanning nine neurotransmitter systems (Neurotransmitter), and cell-type abundance maps derived from the cortical single-nucleus RNA sequencing data comprising 24 neuronal and non-neuronal cell classes (Cell). Dot size represents the strength of the spatial correlation, with small dots indicating nominal significance (P_Spin_ < 0.05) and larger dots indicating associations surviving false discovery rate (FDR) correction (P_FDR-Spin_ < 0.05).

In the ABCD cohort, significant spatial co-localisations were observed primarily at the level of large-scale functional networks. The limbic network showed significant positive co-localisation across disorders (7/10; *r* = 0.33–0.58, P_Spin_ < 0.05), with atypical anorexia nervosa and attention-deficit/hyperactivity disorder remaining significant after FDR correction. Across neurotransmitter systems, dopamine D2 receptor maps showed the greatest number of nominal spatial associations (5/10; *r* = 0.35–0.41, P_Spin_ < 0.05), implicating dopaminergic signalling in deprivation-related brain organisation (Figure 7).

Within the UKB cohort, psychiatric disorders showed no consistent enrichment in canonical functional networks. In contrast, neurological disorders exhibited subcortical network enrichment (5/13; *r* = 0.35–0.74, P_Spin_ < 0.001), with multiple sclerosis and epilepsy remaining significant after FDR correction. At the molecular level, dopamine D2 receptor maps showed spatial associations with psychiatric (7/9; *r* = 0.38–0.54, P_Spin_ < 0.05) and neurological disorders (5/13; *r* = 0.39–0.53, P_Spin_ < 0.05). In addition, the serotonin 5-HT4 receptor map was associated with psychiatric disorders (5/9; *r* = 0.36–0.66, P_Spin_ < 0.05), whereas the norepinephrine transporter map was co-localised with neurological disorders (5/13; *r* = 0.33–0.43, P_Spin_ < 0.05). At the cellular level, psychiatric disorders were most consistently associated with layer 5/6 near-projecting (L5/6 NP) excitatory neurons (6/9; *r* = 0.36–0.63, P_Spin_ < 0.05) and LAMP5-LHX6 inhibitory interneurons (5/9; *r* = 0.36–0.49, P_Spin_ < 0.05). Neurological diseases showed a similar cellular signature, with L5/6 NP neurons (5/13; *r* = 0.38–0.67, P_Spin_ < 0.05), layer 5 intratelencephalic (L5 IT) neurons (5/13; *r* = 0.35–0.64, P_Spin_ < 0.05), and layer 2/3 intratelencephalic (L2/3 IT) neurons (4/13; *r* = 0.39–0.48, P_Spin_ < 0.05) (Figure 7).

Overall, these findings suggest that deprivation preferentially affects limbic systems involved in emotional and motivational processing during neurodevelopment, with modest evidence implicating dopaminergic signalling. In adulthood, deprivation-related mediation patterns become more widely distributed across cortical systems in psychiatric disorders but remain preferentially localised to subcortical circuitry in neurological diseases. At the cellular and molecular levels, these patterns converge on inhibitory interneurons and corticocortical excitatory neuronal populations, together with dopaminergic, serotonergic, and noradrenergic neurotransmitter systems.

### The effects of neighbourhood deprivation on neuroanatomy and neuropsychiatric disease are consistent across cohorts

To evaluate the generalisability of the relationships between deprivation, regional brain volume loss, and neuropsychiatric disease, we first compared the total effects observed in UKB-England with those obtained in two independent cohorts: the developmental ABCD cohort and the geographically distinct UKB-Scotland cohort. In the UKB-England versus ABCD comparison, effect sizes for the nine phenotypically matched disorders were correlated (*r* = 0.63, P = 0.06). Disorders that showed the largest deprivation-related effects in UKB-England were among the strongest associations in ABCD, including cannabis use disorder, tobacco use disorder, and psychotic symptoms. Likewise, the comparison between UKB-England and UKB-Scotland demonstrated substantial replication of total effects across 36 shared diagnoses (*r* = 0.82, P = 8.35 × 10 ^-10^) (Extended Data Figure 2).

We then compared the associations between neighbourhood deprivation and brain structure across the three independent cohorts (HBCD, ABCD and UKB) by examining *a* path coefficient linking deprivation to regional brain measures. Because only six subcortical regions are available in the HBCD dataset, we restricted our analyses to cortical regions. The spatial pattern of deprivation-related effects showed moderate agreement between HBCD and ABCD (r = 0.36, P_Spin_ = 0.04). In contrast, the correspondence between UKB and ABCD (r = 0.17, P_Spin_ = 0.33) and between UKB and HBCD (r = 0.00, P_Spin_ = 0.99) was not statistically significant. These findings suggest that the cortical alterations associated with neighbourhood deprivation in development remain largely consistent into later childhood but differ from associations present in late adulthood (Extended Data Figure 3).

Finally, we examined indirect effects linking neighbourhood deprivation to disease risk through neuroanatomical pathways across cohorts, including five disorders in ABCD and four diseases in UKB-Scotland. Across the nine conditions and two anatomical parcellations, 16 of 18 spatial correlations were positive. In comparisons between UKB-England and ABCD, tobacco use disorder showed a trend-level replication in the cortex (*r* = 0.31, P_Spin_ = 0.082), while obsessive-compulsive disorder (*r* = 0.63, P = 0.069), specific phobia (*r* = 0.59, *P* = 0.096), and sleep problems (*r* = 0.50, P = 0.070) showed trend-level replications in the subcortex, although none survived FDR correction. In comparisons between UKB-England and UKB-Scotland, significant replications were observed for depressive episodes in the subcortex (*r* = 0.81, P_FDR_ = 0.048). In addition, migraine (*r* = 0.46, P_FDR-Spin_ = 0.048) and upper-limb mononeuropathies showed convergent effects in the cortex (*r* = 0.58, P_FDR-Spin_ = 0.018). Together, these cross-cohort analyses indicate that indirect effects linking neighbourhood deprivation to disease risk through neuroanatomical pathways are consistent across independent populations (Extended Data Figure 4).

### Sensitivity analysis

A key consideration in mediation analysis is the assumption of a temporal sequence among variables [30]. In our case, the model posits a deprivation → brain → disease pathway. To better match this temporal ordering in ABCD, we restricted analyses to baseline MRI data and excluded participants with any baseline or historical diagnoses, ensuring that all individuals were healthy at the time of imaging and could develop diagnoses only in later follow-ups. This had minimal impact on sample size because ABCD is a young cohort and most conditions arise only during later follow-up. In contrast, applying the same criterion in the UKB would have substantially reduced the number of cases and excluded many diagnoses, as UKB participants are aged 40–70 and many have already received a clinical diagnosis prior to MRI. For this reason, the primary UKB analyses included all available cases, regardless of diagnosis time. Yet, as a sensitivity analysis, we repeated the UKB-England analyses using only participants whose diagnoses occurred after the imaging date. Results were significantly concordant across all conditions (P_FDR-Spin_ < 0.001), except mononeuropathy of upper limb, which may reflect reduced statistical power due to a smaller sample size (Extended Data Figure 5). Overall, these findings support the robustness of the proposed model in preserving the intended temporal ordering.

Another potential concern is fine-scale population stratification arising from genetic relatedness among participants [31], which could bias estimates of the association between neighbourhood deprivation and brain volume. To assess the robustness of our findings to this possibility, we re-estimated path *a* (deprivation → brain volume) using a kinship-adjusted linear mixed model that accounts for covariance among genetically related individuals via a genetic relationship matrix (GRM) [32] (Methods). Effect estimates were highly concordant with those from the primary analyses across all three cohorts (HBCD, ABCD, and UKB; all P_Spin_ < 0.001), indicating that the deprivation–brain associations were robust to adjustment for population stratification (Extended Data Figure 6).

We next addressed passive gene–environment correlation, whereby biological parents transmit both genetic liability and the rearing environments associated with deprivation. Taking adoption status into account could partly disentangle these influences [33]. The UKB–England sample included 621 adopted participants, which was insufficient for stratified mediation analyses because most individual diagnoses would have had few cases after dividing the sample by adoption status. We therefore focused on path *a* (i.e., deprivation → brain) by extending the primary regression models to include adoption status and an IMD × adoption interaction term. The interaction was not significant in any region (all P_FDR_> 0.05), indicating similar associations between adopted and non-adopted individuals (Supplementary Table 10). Although this analysis cannot rule out genetic confounding, it supports the assertion that the rearing environment is not the primary explanation for the observed associations.

## Discussion

Using three large population-based cohorts spanning the earliest stages of life through late adulthood, we demonstrate that neighbourhood deprivation is consistently associated with increased disease risk, with individual lifestyle factors accounting for only a small proportion of these associations. Across independent cohorts, greater neighbourhood deprivation was also associated with widespread reductions in cortical and subcortical brain volume, with regional brain structure consistently mediating the association between deprivation and neuropsychiatric disorders. Deprivation-related structural alterations preferentially involved brain regions intrinsically vulnerable to neuropsychiatric disorders, while spatial decoding implicated dopaminergic and serotonergic neurotransmitter systems together with specific excitatory and inhibitory neuronal cell classes. Our findings were consistently replicated across independent cohorts.

Although previous epidemiological studies have linked socioeconomic disadvantage with individual conditions such as mood disorders [8] and dementia [9], most investigations have focused on a limited number of diseases or a single developmental stage. Our findings suggest that neighbourhood deprivation is not disorder-specific but instead represents a general environmental risk factor that increases vulnerability across multiple diagnostic domains throughout the life span. This broad pattern of associations highlights neighbourhood deprivation as an important public health target whose impact extends well beyond individual diseases.

Another novel contribution of this study was to test whether lifestyle or broader environmental factors link deprivation to neuropsychiatric disorders. If lifestyle behaviours were the primary pathway linking neighbourhood deprivation to disease, little of the IMD’s effect should remain after accounting for these behaviours. Instead, adjustment for alcohol intake, smoking, diet, sleep, sun exposure, and physical activity modestly reduced the estimated IMD, with nearly all associations remaining statistically significant. These findings support the view that broader characteristics of the residential environment—including chronic psychosocial stress and unequal access to healthcare, education, and other community resources—may independently contribute to disease risk [34, 35]. Consequently, reducing health inequalities is likely to require policies that address ecological and societal conditions alongside interventions targeting individual behaviours.

Another important observation, not explored in prior work, is the consistent deprivation-related reduction in regional brain volume across three cohorts spanning the life. Although neonatal associations were not statistically significant, likely reflecting the limited sample size and the relatively short duration of environmental exposure, the direction of effects was already apparent during the neonatal period and became widespread during later childhood and adulthood. This developmental pattern is compatible with theories proposing that socioeconomic adversity exerts cumulative effects on brain maturation [36, 37]. These findings suggest that interventions aimed at reducing socioeconomic disadvantage may have the greatest impact when implemented early in development, before deprivation-related neuroanatomical differences become more widespread.

We also showed that regional brain volume partially mediated the association between neighbourhood deprivation and disease risk. Across both ABCD and UKB, mediation effects showed striking consistency in direction. In nearly all disorders, greater deprivation was associated with lower regional brain volume, which in turn was associated with higher disease risk. One possible explanation for the current findings is the allostatic load framework [38, 39], which proposes that chronic socioeconomic adversity produces widespread physiological dysregulation through repeated activation of stress-response systems. Persistent alterations in glucocorticoid signalling [40] and inflammation [41] may influence neural plasticity and brain development, ultimately contributing to regional brain atrophy [42]. Nevertheless, the precise biological pathways linking allostatic load to structural brain alterations remain to be established and should be investigated in future longitudinal and mechanistic studies.

Importantly, we indicated that regions exhibiting stronger mediation effects also showed greater disease-related volume loss in independent cohorts. These observations support the concept of selective regional vulnerability, whereby stressors preferentially affect regions with heightened susceptibility to pathological processes [43, 44]. Consequently, exposure to disadvantaged neighbourhood environments may increase the population burden of neuropsychiatric disorders by shifting intrinsically vulnerable brain regions closer to pathological thresholds, thereby reducing resilience to subsequent genetic, biological, or environmental insults. Our findings therefore highlight the neurobiological value of reducing neighbourhood deprivation, suggesting that improvements in neighbourhood conditions may help preserve brain resilience and reduce the burden of neuropsychiatric disorders.

Spatial decoding analyses placed these deprivation-related brain alterations within a broader neurobiological framework. During childhood and adolescence, mediation patterns were most consistently enriched within the limbic network and showed modest spatial correspondence with dopamine D2 receptor distribution. The limbic system plays a central role in emotional processing, reward learning, and social behaviour [45, 46]. Dopamine D2 receptors are highly expressed within mesolimbic and corticostriatal circuits, regulating reward processing, reinforcement learning, and behavioural flexibility [47, 48]. Because these circuits exhibit high developmental plasticity [49], they may be particularly susceptible to socioeconomic adversity, which could alter the maturation of emotional and motivational systems.

In adulthood, deprivation-related brain changes showed common and distinct neurobiological signatures for psychiatric and neurological disorders. For psychiatric disorders, spatial associations were most consistently observed with dopaminergic and serotonergic neurotransmitter systems. Building on the developmental involvement of dopaminergic circuits, the additional enrichment of serotonergic systems suggests that the neurobiological consequences of deprivation extend beyond reward-related processes. Serotonin plays a central role in affective processing, emotional regulation, and stress adaptation [50, 51], and alterations in serotonergic signalling have long been implicated in mood, anxiety, and related psychiatric disorders [52, 53]. At the cellular level, deprivation-related brain maps were enriched for layer 5/6 near-projecting (L5/6 NP) excitatory neurons and LAMP5-LHX6 interneurons. Enrichment in both excitatory and inhibitory cell populations provides a plausible mechanistic route by which deprivation could disrupt the excitation–inhibition balance, a feature that has been proposed as transdiagnostic across several psychiatric disorders [54, 55].

In contrast, neurological diseases remained preferentially localised to subcortical systems, consistent with the prominent involvement of subcortical structures in the pathology of many neurological diseases. At the molecular level, these patterns were associated with distributions of the dopamine D2 receptor and the norepinephrine transporter (NET). Whereas dopamine plays a central role in basal ganglia function and motor control [56], noradrenergic signalling regulates arousal, attention, autonomic function [57], and neuroinflammatory responses [58], processes that are often disrupted in neurological disease. Cell-type enrichment analyses further implicated L5/6 near-projecting (NP), L5 intratelencephalic (IT), and L2/3 IT neurons, indicating a preferential association with excitatory projection neurons spanning deep and superficial cortical layers. In particular, the involvement of L5 IT neurons, which contribute to corticostriatal as well as corticocortical projections [59], may complement the prominent subcortical localisation of neurological dysfunctions. Although spatial decoding cannot determine causal mechanisms, it generates biologically grounded hypotheses about pathways through which socioeconomic disadvantage may confer neuropsychiatric risk and points to systems that warrant future targeted investigation.

One notable aspect of our findings is their reproducibility across independent populations. Associations between deprivation and disease replicated across geographically distinct UKB subgroups and showed similar trends in the developmentally independent ABCD cohort. Likewise, mediation patterns demonstrated significant spatial correspondence across cohorts despite differences in participant age, imaging protocols, healthcare systems, diagnostic classifications, and measures of neighbourhood deprivation. Such consistency strengthens confidence that the observed relationships reflect general biological principles rather than cohort-specific artefacts or methodological biases.

Despite these strengths, several limitations should be considered. First, the observational nature of the study precludes definitive causal inference. Although our mediation framework was supported by sensitivity analyses addressing temporal ordering, population stratification, and passive gene–environment correlation, unmeasured confounding and bidirectional relationships cannot be fully ruled out [60]. Second, while we examined neighbourhood deprivation using the ADI and IMD, these measures primarily capture the socioeconomic dimensions of deprivation. Future research is needed to investigate how environmental deprivation may influence disease risk through its effects on the brain. Third, the analyses were restricted to participants of European ancestry to minimise population stratification and maximise comparability across cohorts, which may limit the generalisability of the findings to other ancestral populations. Fourth, substantial sample-size differences between HBCD, ABCD, and UKB could bias cross-cohort contrasts by amplifying noise in smaller samples and stabilising estimates in larger ones.

In summary, the cross-cohort convergence of epidemiological, neuroanatomical, and molecular evidence indicates that neighbourhood deprivation is a broad environmental determinant of neuropsychiatric vulnerability across development. Reducing socioeconomic disadvantage through sustained investment in components of the ADI and IMD—such as housing, education, healthcare access, and community infrastructure—may help preserve brain resilience and lower the population burden of disease.

## Methods

### Datasets

#### HEALthy Brain and Child Development (HBCD) Study

The HBCD is an ongoing, prospective longitudinal cohort designed to recruit over 7,000 mother– infant dyads across 27 sites in the United States and follow participants from pregnancy through 10 years of age. The study collects repeated multimodal assessments to characterise neurodevelopment, including structural, functional, diffusion, quantitative MRI, magnetic resonance spectroscopy, EEG, biospecimens, and comprehensive behavioural and environmental phenotyping. Behavioural assessments encompass cognitive and language development, motor development, temperament, emotional and behavioural functioning, self-regulation, caregiver– child interactions, sleep, physical health and growth, nutrition, family environment, parental mental health, substance use, stress, social support, adverse childhood experiences, and neighbourhood and environmental exposures. MRI assessments are performed at four longitudinal time points beginning in early infancy (0–1 month of age; Visit 2). In the present study, we analysed baseline MRI data acquired at Visit 2, corresponding to the first postnatal neuroimaging assessment.

#### Adolescent Brain Cognitive Development (ABCD) Study

The ABCD Study is an ongoing longitudinal, population-based study of brain development and child health in the United States. Between 2016 and 2018, the study recruited 11,878 children aged 9–10 years from 21 research sites distributed across the country. Participants underwent comprehensive assessments of cognitive function, mental and physical health, environmental exposures, and socioeconomic circumstances, together with multimodal neuroimaging and genetic data collection. Follow-up assessments are conducted annually, with major biennial visits including repeated neuroimaging acquisitions. For this study, we used data from the baseline imaging assessment together with information from the *Kiddie Schedule for Affective Disorders and Schizophrenia (KSADS)*. We identified *416* healthy participants, all of whom had neuroimaging data available at baseline. Additional details regarding case and control definitions are provided in the *Outcomes* section.

#### UK Biobank (UKB)

The UKB is a large, population-based prospective cohort study that recruited approximately 500,000 participants aged >40 years between 2006 and 2010 across the United Kingdom. Participants underwent extensive phenotypic assessments, including demographic, socioeconomic, lifestyle, cognitive, physical, and health-related evaluations, and provided biological samples for genetic analyses. Beginning in 2014, a subset of participants also underwent multimodal neuroimaging assessments, including structural magnetic resonance imaging (MRI). The UKB provides longitudinal follow-up through linkage to electronic health records, including primary care, hospital inpatient, cancer registry, and death registry data, enabling comprehensive ascertainment of disease outcomes. For the present study, we used data from participants’ first imaging visit together with information on *International Classification of Diseases (ICD)* diagnoses. The study included *225,877* healthy controls, of whom *39,789* had neuroimaging data available. Additional details regarding case and control definitions are provided in the *Outcomes* section.

### Exposure

In the HBCD and ABCD Studies, socioeconomic disadvantage was measured using the *Area Deprivation Index (ADI)*, a validated composite measure of neighbourhood socioeconomic disadvantage. The ADI is derived from 18 census-based socioeconomic indicators from the 2011– 2015 American Community Survey and is available as both a weighted score and a national percentile [61]. Census-tract-level ADI values were assigned to participants’ primary, secondary, and tertiary residential addresses at baseline, and the primary ones were used in all analyses. A higher ADI score reflects living in a relatively more deprived area.

In the UKB, socioeconomic disadvantage was quantified using the *Index of Multiple Deprivation (IMD)* as a continuous exposure measure. The IMD is the official area-level measure of multiple deprivation in the UK, calculated by the UK government’s Office for National Statistics. It combines seven domains: income, employment, education, health and disability, crime, barriers to housing and services, and living environment. Compared with simpler socioeconomic indices, such as the Townsend Deprivation Index, the IMD provides a multidimensional assessment of neighbourhood disadvantage. Because deprivation indices are derived separately for England, Scotland, and Wales using different methodologies, UKB provides country-specific IMD scores without harmonisation across nations. A higher IMD score indicates that a participant resides in a relatively more deprived area.

### Mediators

In HBCD, T1-weighted (T1w) and T2-weighted (T2w) structural MRI images underwent denoising, bias-field correction, and spatial normalisation to the age-appropriate MNI Infant template (0–4.5 years). The images were then transformed to the MNI152 template to ensure compatibility with older cohorts. Due to incomplete neonatal myelination and reduced grey– white matter contrast on T1w images, cortical surface reconstruction was performed using T2w images. This was carried out within the fMRIPrep Lifespan [23] using the Surface-based Melbourne Children’s Regional Infant Brain (M-CRIB-S) method [62]. Specifically, a modified MCRIBReconAll workflow was implemented, with BIBSNet-derived brain segmentations provided as external inputs to guide reconstruction [63]. Structural image quality was assessed using MRIQC [64]. Full details of the HBCD MRI preprocessing procedures are reported in [65].

For ABCD and UKB, we downloaded minimally processed T1- and T2-FLAIR-weighted images. Images were processed with FreeSurfer (v6.0.1) [66, 67], using T2-FLAIR-weighted images, when available, to improve pial surface reconstruction. The recon-all pipeline included bias field correction, stereotaxic registration, intensity normalisation, skull stripping, and white matter segmentation. A triangular surface tessellation was used to fit a deformable mesh to the white matter volume, generating grey–white and pial surfaces with >160,000 corresponding vertices registered to fsaverage standard space. Cortical surfaces were reconstructed for each participant and aligned to fsaverage using FreeSurfer’s surface-based registration. Subcortical structures were automatically segmented and labelled using FreeSurfer’s volumetric segmentation procedure.

Mediators in our analysis comprised 34 bilaterally averaged cortical regions [24] across HBCD, ABCD, and UKB, along with 9 subcortical regions (Accumbens-area, Amygdala, Brain-Stem, Caudate, Cerebellum-Cortex, Hippocampus, Pallidum, Putamen, and Thalamus-Proper), of which 6 were available in HBCD (Amygdala, Brain-Stem, Caudate, Cerebellum-Cortex, Hippocampus, and Thalamus).

### Outcomes

For both UKB and ABCD, analyses were restricted to diagnostic outcomes with at least 50 cases. Participants with no recorded diagnosis of any of the diseases included in the analyses were classified as healthy controls.

The ABCD study used the computerised KSADS-5, a DSM-5–based structured diagnostic interview with established reliability in youth populations, delivered in clinician-, parent-, and youth-report formats. The computerised parent and youth versions show high concordance with clinician-administered KSADS-5 (88–96% diagnostic agreement), supporting their use in large-scale data collection. For ABCD, the instrument was adapted in collaboration with its developers through item-wording revisions, algorithm adjustments to permit past-year diagnoses, modularisation for flexible administration, and Spanish translation. At baseline and major biennial imaging visits, parents complete nearly all KSADS-5 modules except enuresis, encopresis, and selective mutism; traumatic experiences are captured within the parent PTSD module. Annual in-person visits repeat the full parent-report KSADS-5, while shorter interim assessments administer only the externalising, psychosis, and eating-disorder modules, reflecting evidence that parent report is most informative for these domains in this age range. Additional details are provided elsewhere [68]. We used 24 diagnostic variables from the parent-reported KSADS-5.

In UKB, outcomes were derived by mapping multiple sources of clinical information to ICD codes. These sources included Primary Care records (Category 3000), ICD-9 and ICD-10 codes from Hospital Inpatient data (Category 2000), ICD-10 codes from Death Register records (Fields 40001 and 40002), and self-reported medical condition codes collected at baseline and subsequent assessment centre visits (Field 20002). We utilised 109 ICD-10 diagnoses recorded in England, comprising 32 disorders from Chapter F (Mental and Behavioural Disorders) and 77 diseases from Chapter G (Diseases of the Nervous System). Chapter F encompasses mood, anxiety, psychotic, developmental, and substance-related disorders, whereas Chapter G includes neurodegenerative diseases, epilepsy, movement disorders, demyelinating diseases, and other neurological conditions.

### Demographic covariates

We included sex, age, age², sex × age and sex × age². These covariates were included to adjust for potential confounding arising from linear and non-linear age effects and sex-by-age interactions.

### Genetic covariates

We accounted for genetic ancestry by including the first 10 ancestry principal components (PCs), estimated using the GENESIS R package [32], as covariates in all analyses. Briefly, pre-processed genotype data from all cohorts were obtained. ABCD and UKB data underwent further processing as described in our previous work [69, 70]. Variants were pruned for linkage disequilibrium (LD) using the SNPRelate package with a correlation-based approach, a 10 Mb sliding window, and an LD threshold of *r*² < 0.1 to retain independent markers. Pairwise kinship coefficients were then estimated from the LD-pruned genotype data using KING [71], with a kinship threshold of 2^-11/2^, corresponding to third-degree relatives. Principal components were subsequently estimated using PC-AiR, which derives ancestry principal components from a mutually unrelated subset of individuals representative of the sample’s ancestral diversity and then projects related individuals onto these ancestry axes. To further minimise population stratification, all analyses were restricted to participants with genetically inferred European ancestry.

For the sensitivity analysis, we constructed a genetic relationship matrix (GRM) to model covariance among individuals. LD-pruned genotype data were used as input to PC-Relate, which estimates pairwise kinship coefficients while accounting for ancestry structure via the PC-AiR principal components. The resulting kinship estimates were converted into a sparse GRM using a threshold of 2^-11/2^. The GRM was then supplied as the random-effects covariance matrix in the GENESIS linear mixed model, ensuring that the deprivation–brain associations were evaluated while explicitly modelling familial relatedness.

### Neuroimaging covariates

Imaging-related covariates were selected according to participant age and the image preprocessing pipeline used in each cohort. Because infant MRI is particularly susceptible to motion artefacts and presents greater preprocessing challenges, more stringent image quality control procedures were applied in the HBCD cohort. Specifically, an extended set of MRIQC-derived quality metrics was used as covariates, comprising T2-weighted contrast-to-noise and signal-to-noise ratio, mean framewise displacement (FD), and the number of high-FD volumes. In the ABCD cohort, head motion was similarly accounted for by adjusting for mean and maximum FD. Because both the ABCD and UKB cohorts underwent cortical reconstruction using FreeSurfer, regression models were also adjusted for the Surface Holes metric, an indicator of structural image quality. Finally, imaging site was accounted for in all cohorts to capture site-related variability and potential scanner-related differences.

### Handling Outliers

To improve the robustness of the analyses, median absolute deviation (MAD) and standard deviation (SD) were calculated for all covariates as well as cortical and subcortical imaging measures. In the HBCD cohort, where infant MRI requires more stringent quality control, observations exceeding **±3 MAD or ±3 SD** from their respective distributions were excluded. In the ABCD and UKB cohorts, outliers were defined using the **±5 MAD or ±5 SD** criterion. This procedure minimises the influence of extreme values arising from factors such as population stratification, poor image quality or preprocessing, and atypical brain morphology, thereby improving the robustness and reliability of the statistical analyses.

### Regression Models

Analyses were implemented in Python using the statsmodels package. Mediation analyses were conducted separately for each exposure–mediator–outcome triplet.

#### 1. Path a: IMD → Mediator

Path a represents the association between IMD (X) and the mediator (M). It was estimated using ordinary least squares regression:

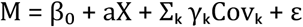

where a represents the association between IMD and the mediator, γₖ represents the regression coefficient for the k^th^ covariate, and ε represents the residual error term.

#### 2. Path b: Mediator → disease outcome

Path b represents the association between the mediator (M) and the binary disease outcome (Y), adjusting for IMD. It was estimated using a binomial generalised linear model with a logit link:

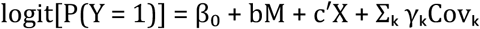

where b represents the association between the mediator and the disease outcome, conditional on IMD and the included covariates.

#### 3. Direct effect (c′): IMD → disease outcome

The direct effect (c′) represents the association between IMD and the disease outcome after adjustment for the mediator. It was obtained as the coefficient for IMD (X) from the same outcome model used to estimate path b:

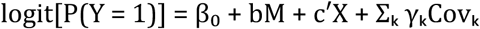

Thus, c′ represents the association between IMD and the disease outcome conditional on the mediator and the included covariates.

#### 4. Indirect effect

The indirect effect was calculated using the product-of-coefficients approach:

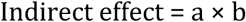

where a represents the association between IMD and the mediator and b represents the association between the mediator and the disease outcome, conditional on IMD and the included covariates.

#### 5. Total effect (c)

The total effect (c) represents the overall association between IMD and the disease outcome before accounting for the mediator. It is estimated using a binomial generalised linear model with a logit link:

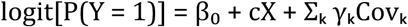

where c represents the association between IMD and the disease outcome without adjustment for the mediator.

### Bootstrap Procedure

To obtain robust estimates of the statistical model, we used a nonparametric bootstrap procedure with 5,000 iterations for each regression analysis. In each iteration, cases were resampled with replacement, while controls were independently resampled with replacement from the predefined control pool. The parameters were re-estimated for each bootstrap sample, and the resulting estimates were retained to derive the empirical distributions for all the paths in the model.

### Statistical Inference

For each parameter distribution, we computed:

● Percentile confidence intervals (2.5th and 97.5th percentiles)
● Two-sided bootstrap p-values, defined as:

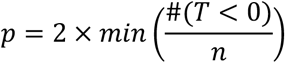

where *T* is the bootstrap distribution of the parameter.

### Lifestyle variables in UKB

Lifestyle measures were derived from the UKB baseline assessment and included diet, sun exposure, physical activity, sleep duration, smoking status, and alcohol intake frequency. Diet was characterised using vegetable intake (Fields 1289 and 1299), fruit intake (1309 and 1319), fish intake (1329 and 1339), processed and unprocessed meat intake (1349, 1359, 1369, 1379, 1389, and 3680), cheese and dairy consumption (1408 and 6144), milk and spread type (1418, 1428, 2654, and 10767), bread and cereal intake and type (1438–1468 and 10776), salt added to food (1478), tea, coffee, and water intake (1488–1528), recent dietary change and dietary variation (1538, 1548, and 10912), and dietary exclusions (10855). Sun exposure was characterised using time spent outdoors in summer and winter (Fields 1050 and 1060), skin colour (1717), tanning ability (1727), childhood sunburn frequency (1737), natural hair colour (1747), facial ageing (1757), use of sun/UV protection (2267), and solarium use (2277). Physical activity was assessed using overall physical activity (Field 22034), sleep duration using self-reported sleep duration (Field 1160), smoking using smoking status (Field 20116), and alcohol consumption using alcohol-intake frequency (Field 1558).

Because diet and sun exposure were each represented by multiple correlated variables, rather than a single measure such as smoking status or alcohol intake frequency, they were summarised by the first principal component using the Principal Component Analysis implementation in scikit-learn (sklearn.decomposition.PCA) after standardising the contributing variables to zero mean and unit variance with sklearn.preprocessing.StandardScaler.

To assess the contribution of lifestyle behaviours to deprivation-associated disease risk, we repeated the primary analyses in the UKB-England cohort after additional adjustment for lifestyle variables. The contribution of lifestyle adjustment was quantified as the percentage change in the IMD odds ratio (OR), calculated as

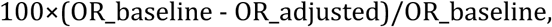

where OR_baseline denotes the OR from the baseline model, and OR_adjusted denotes the OR after additional adjustment for lifestyle factors.

### Preprocessing of lifestyle variables in UKB

Variable-specific data cleaning and re-coding were performed according to the built-in FMRIB configuration of FUNPACK (FMRIB UK Biobank Normalisation, Parsing and Cleaning Kit) (https://pages.fmrib.ox.ac.uk/fsl/funpack/demo.html). These procedures include replacing predefined non-informative response codes with missing values, data-field-specific cleaning and reformatting, recoding categorical values, and, where applicable, replacing missing values in dependent variables based on their parent data fields. Following this preprocessing, variables with more than 5% missing observations in the analytical sample were excluded from subsequent analyses.

### Case-control maps

To generate case–control maps for each disease, we first regressed out the effects of genetic, demographic, and neuroimaging confounders from the regional brain volumes. The corresponding neighbourhood deprivation measure (ADI in ABCD and IMD in UKB-Scotland) was also included as a covariate to ensure that the resulting case–control maps were independent of neighbourhood deprivation. We then computed t-values to quantify regional volumetric differences between cases and controls, with negative values indicating lower regional volumes in cases than in controls. We quantified the spatial correspondence between disease-related *t*-maps and deprivation-related mediation β-maps. These comparisons were performed irrespective of whether the mediation or case–control effects were statistically significant. We also excluded disease categories classified as “other” because they are not uniformly defined and comprise heterogeneous conditions, making them unsuitable for reliable cross-cohort comparisons.

### Biological Decoding

#### Canonical functional networks [26]

The authors identified seven intrinsic functional networks by clustering resting-state functional connectivity patterns in a large cohort of healthy individuals, capturing the principal functional organisation of the human cerebral cortex. These networks comprise the visual, somatomotor, dorsal attention, ventral attention (salience), limbic, frontoparietal control, and default mode networks, each supporting distinct cognitive and behavioural functions. The visual and somatomotor networks are primarily involved in sensory processing and motor control, whereas the dorsal and ventral attention networks support externally directed attention, salience detection, and cognitive reorienting. The frontoparietal control network is associated with executive control and flexible goal-directed behaviour, while the default mode network is implicated in internally directed cognition, including autobiographical memory, self-referential thought, and future planning. The limbic network contributes predominantly to emotion, reward processing, and memory.

#### Neurotransmitter systems [27]

This atlas integrates PET imaging data from healthy individuals to generate cortical maps that represent the spatial distribution of 19 molecular targets across nine major neurotransmitter systems. These include the serotonin receptors and transporter (5-HT1A, 5-HT1B, 5-HT2A, 5-HT4, 5-HT6, and 5-HTT), the cholinergic markers (α4β2 nicotinic acetylcholine receptor, M1 muscarinic receptor, and vesicular acetylcholine transporter [VAChT]), the dopaminergic markers (D1 receptor, D2 receptor, and dopamine transporter [DAT]), the GABAa receptor, histamine H3 receptor, cannabinoid CB1 receptor, μ-opioid receptor (MOR), norepinephrine transporter (NET), and the glutamatergic markers (N-methyl-D-aspartate receptor [NMDAR] and metabotropic glutamate receptor 5 [mGluR5]). These neurotransmitter systems provide a molecular framework for interpreting macroscale neuroimaging findings.

#### Cellular architecture [29]

Authors generated an atlas of cortical cell-type abundance using single-nucleus RNA sequencing (snRNA-seq) across eight neocortical regions, including the primary motor cortex (M1), primary somatosensory cortex (S1), primary auditory cortex (A1), primary visual cortex (V1), dorsolateral prefrontal cortex (DFC), anterior cingulate cortex (ACC), middle temporal gyrus (MTG), and angular gyrus (AnG). These regions span the rostral-to-caudal extent of the cortex and capture major variations in cortical cytoarchitecture. The resulting atlas comprised 24 cortical cell classes with distinct developmental origins, laminar specialisation, morphology, electrophysiological properties, and projection patterns, including nine GABAergic interneuron classes (PAX6-expressing, SNCG-like, vasoactive intestinal peptide [VIP]-expressing, LAMP5-expressing, LAMP5/LHX6-expressing, chandelier, parvalbumin [PVALB]-expressing, somatostatin [SST]/CHODL-expressing, and somatostatin [SST]-expressing interneurons), nine glutamatergic excitatory neuron classes comprising intratelencephalic-projecting neurons from layers 2/3, 4, 5, and 6, including a layer 6 Car3-like subtype, together with extratelencephalic-projecting layer 5 neurons, near-projecting layer 5/6 neurons, corticothalamic-projecting layer 6 neurons, and layer 6b neurons, and six non-neuronal cell classes (astrocytes, endothelial cells, microglia/perivascular macrophages, oligodendrocytes, oligodendrocyte precursor cells, and vascular leptomeningeal cells).

To obtain cell-type maps in the Desikan–Killiany atlas, we followed a recent work that deconvolves bulk gene expression data from the Allen Human Brain Atlas (AHBA) and derives cellular signatures from Jorstad’s work [72]. Further details are available at https://github.com/XihanZhang/human-cellular-func-con/tree/main/cell_maps.

## Data Availability

The participant-level data analysed in this study are controlled-access and cannot be redistributed by the authors. ABCD Study and HBCD Study data are available to eligible researchers through the NIH Brain Development Cohorts Data Hub (https://www.nbdc-datahub.org/), subject to approval of a Data Use Certification and completion of the required training. UK Biobank data are available to eligible researchers through the UK Biobank access process (https://www.ukbiobank.ac.uk/use-our-data/apply-for-access/).

## Ethics Statement

This research was conducted using the UK Biobank Resource under application number 20904 and utilised anonymised data under the existing Research Tissue Bank (RTB) approval.

## Author contribution

Conceptualisation: A. E., R. B., and T. R. Methodology: All authors. Investigation: A. E. Visualisation: A. E. Supervision: R. B., V. W., and T. R. Writing—original draft: A. E. Writing—review and editing: All authors.

## Code availability

All code written for this study is publicly available (https://github.com/amir-ebneabbasi/Deprivation-Brain-Health).

---

**Extended Data Figure 1.**
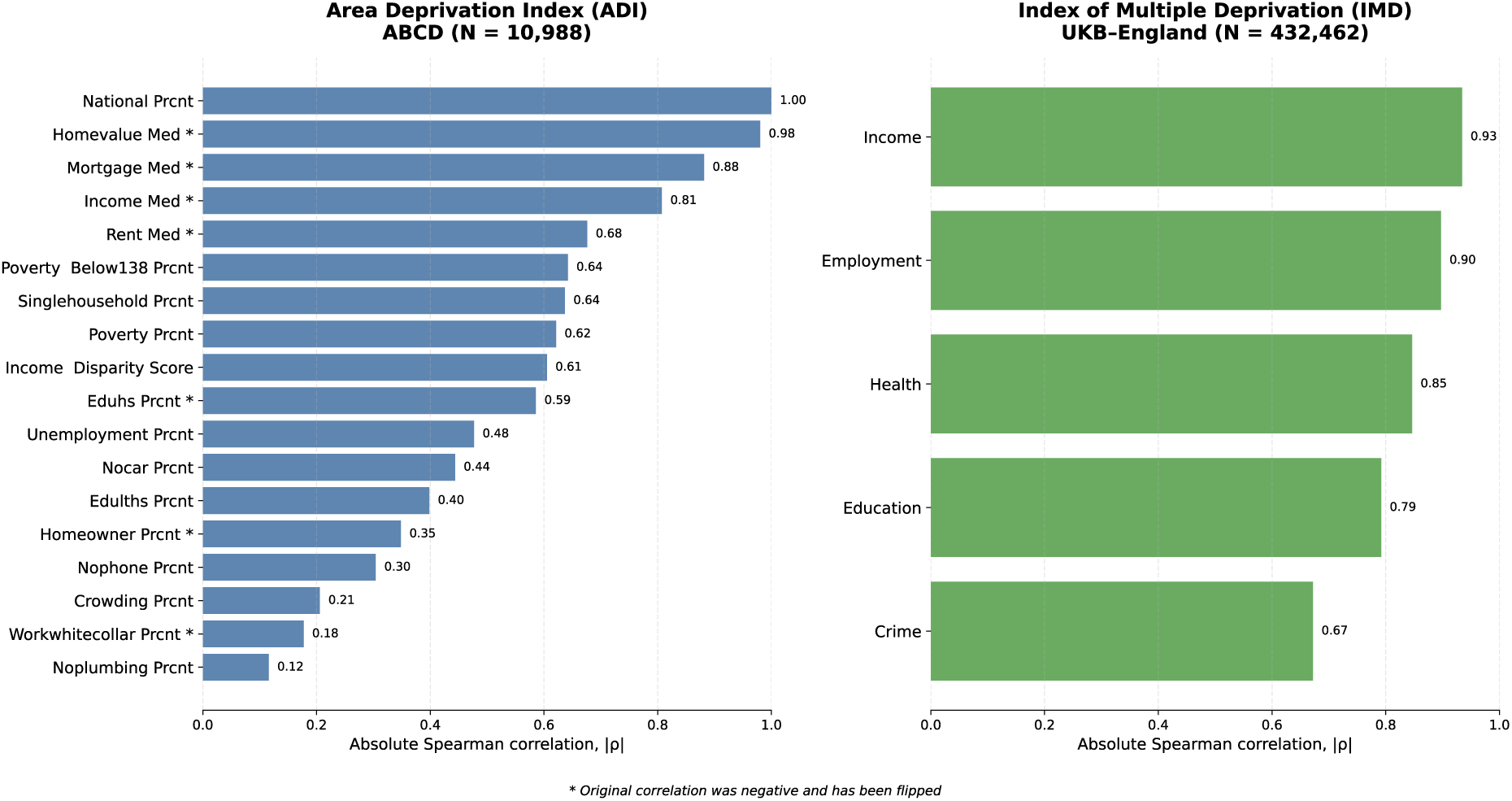
Correlations between composite neighbourhood deprivation indices and their constituent socioeconomic indicators.

**Extended Data Figure 2.**
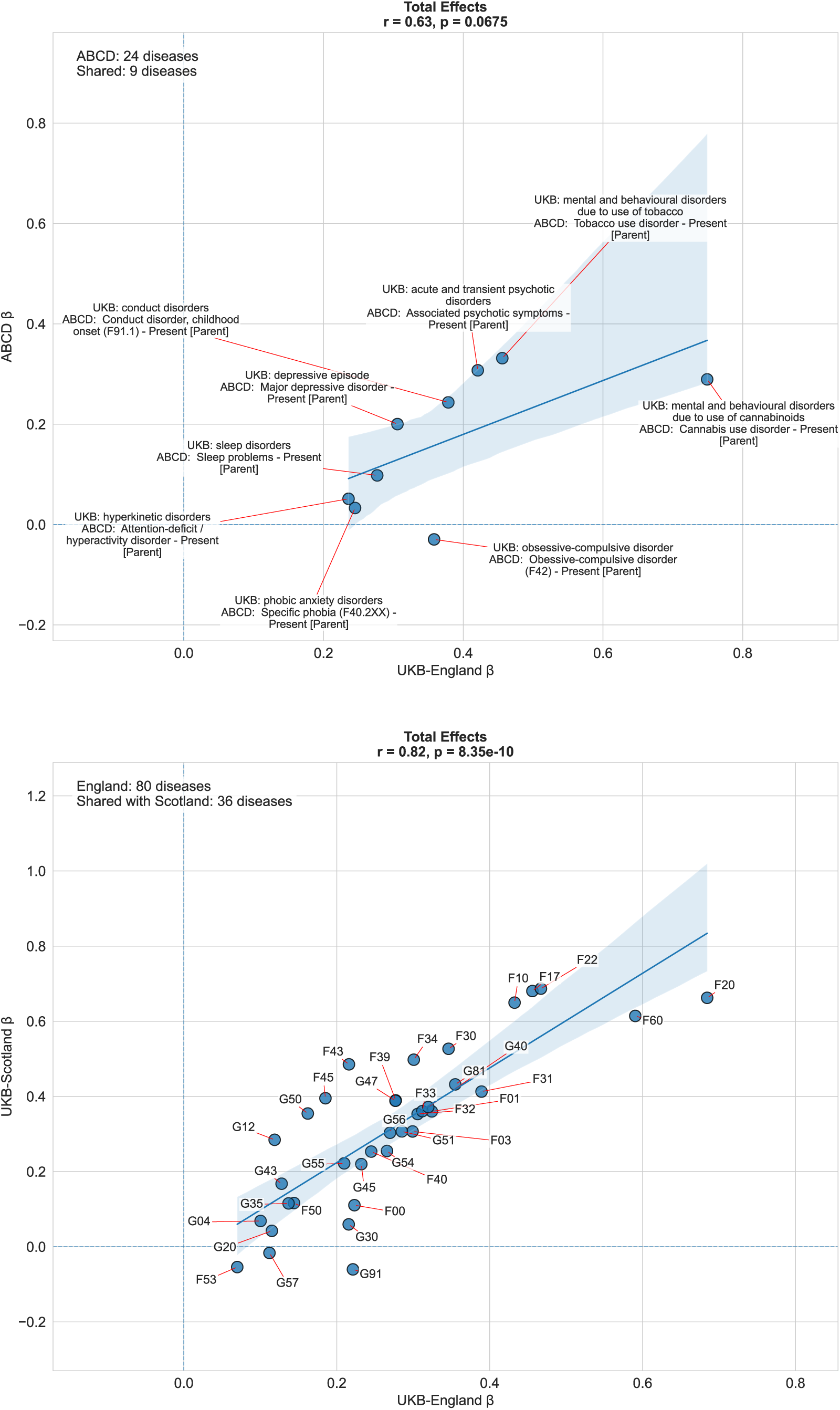
Cross-cohort comparison of total deprivation effects on disease outcomes.

**Extended Data Figure 3.**
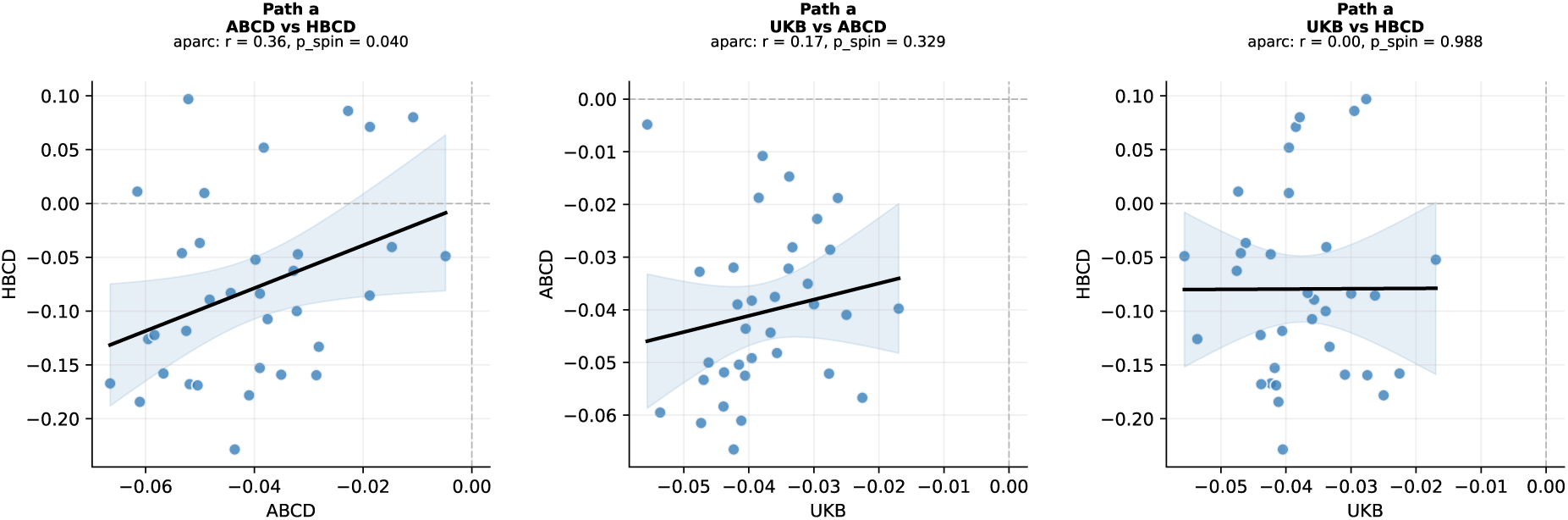
Cross-cohort comparison of deprivation effects on brain volume.

**Extended Data Figure 4.**
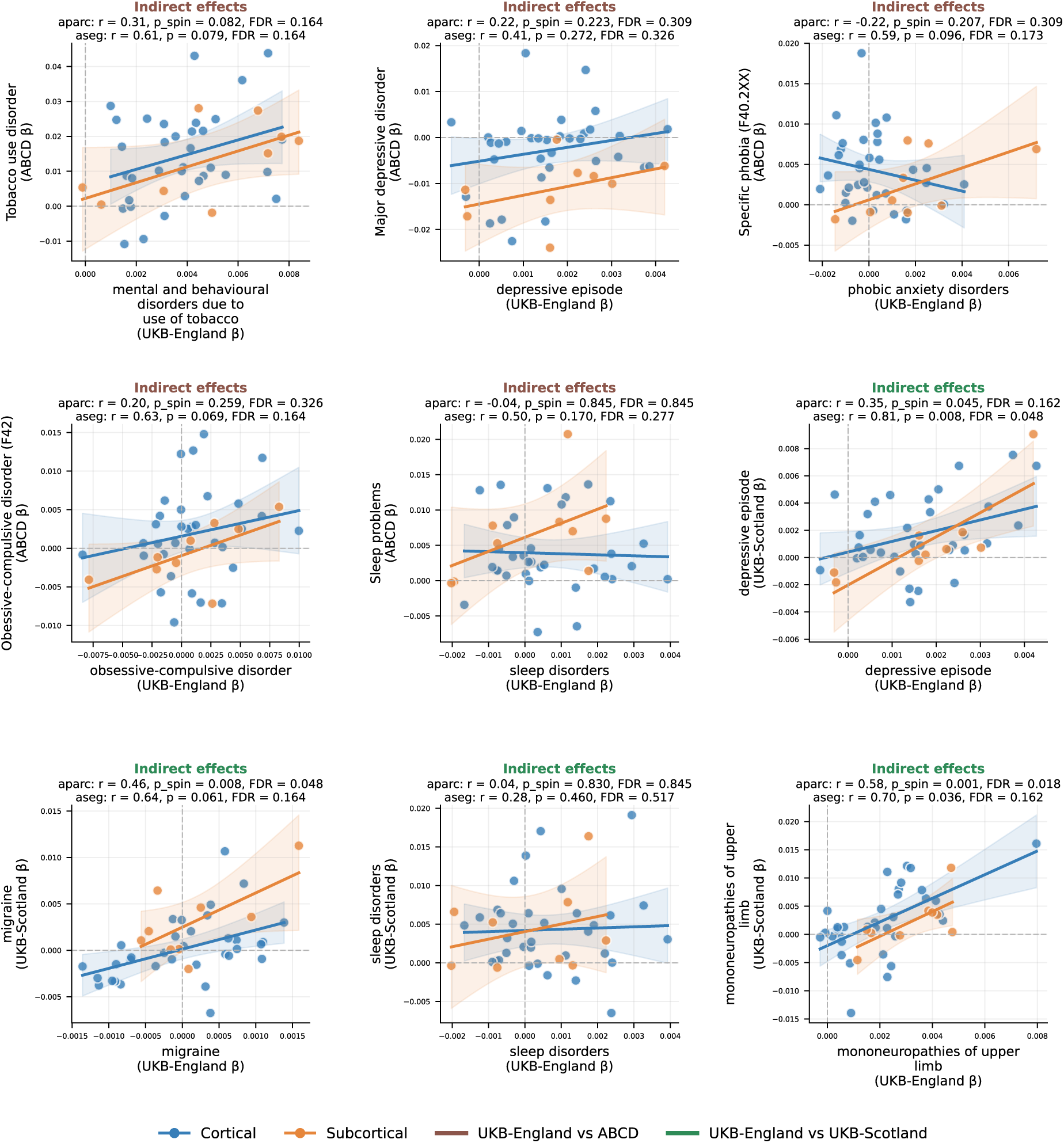
Cross-cohort comparison of indirect deprivation effects on disease outcomes.

**Extended Data Figure 5.**
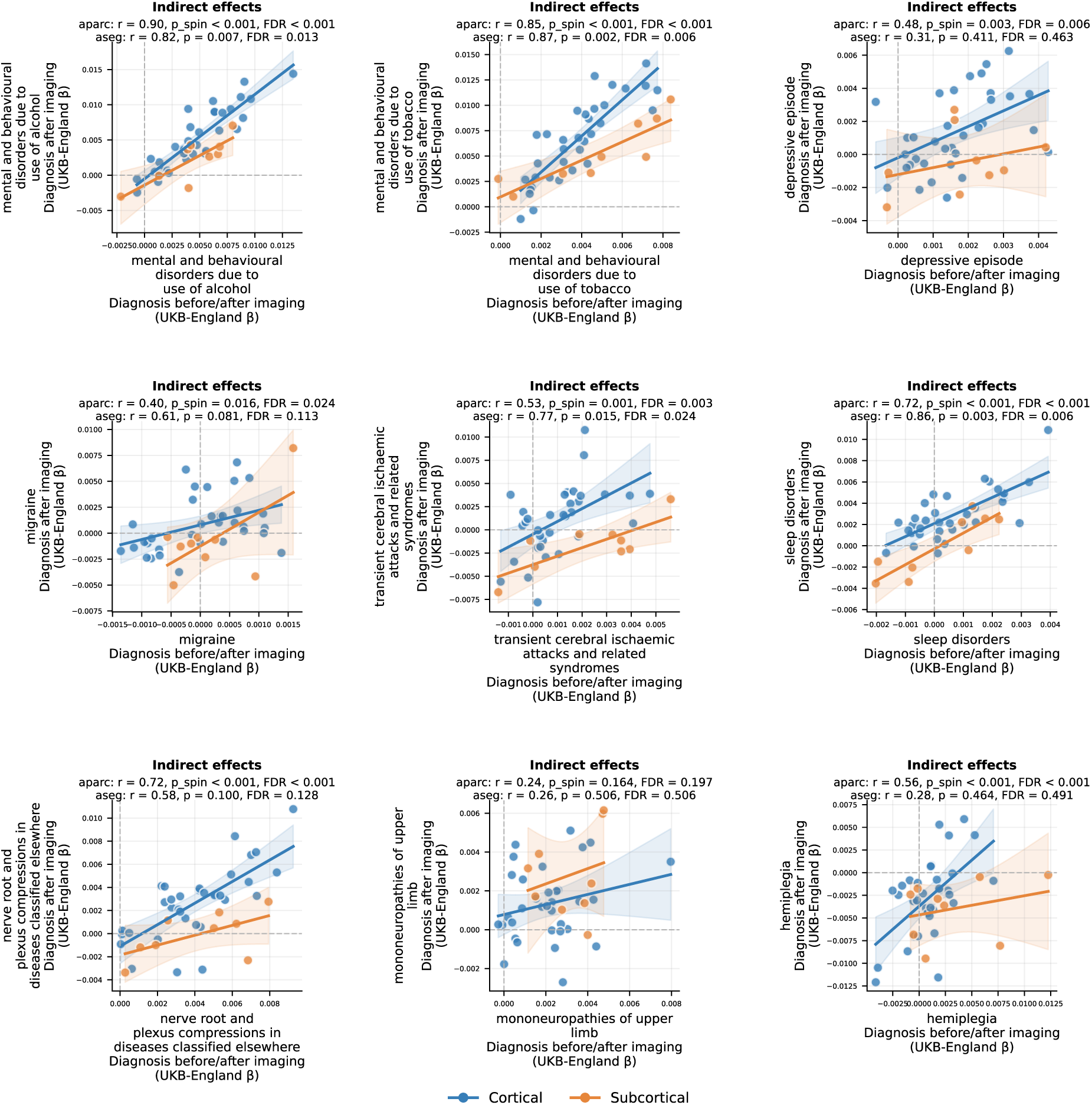
Comparison of indirect deprivation effects on diseases across different dates of diagnosis.

**Extended Data Figure 6.**
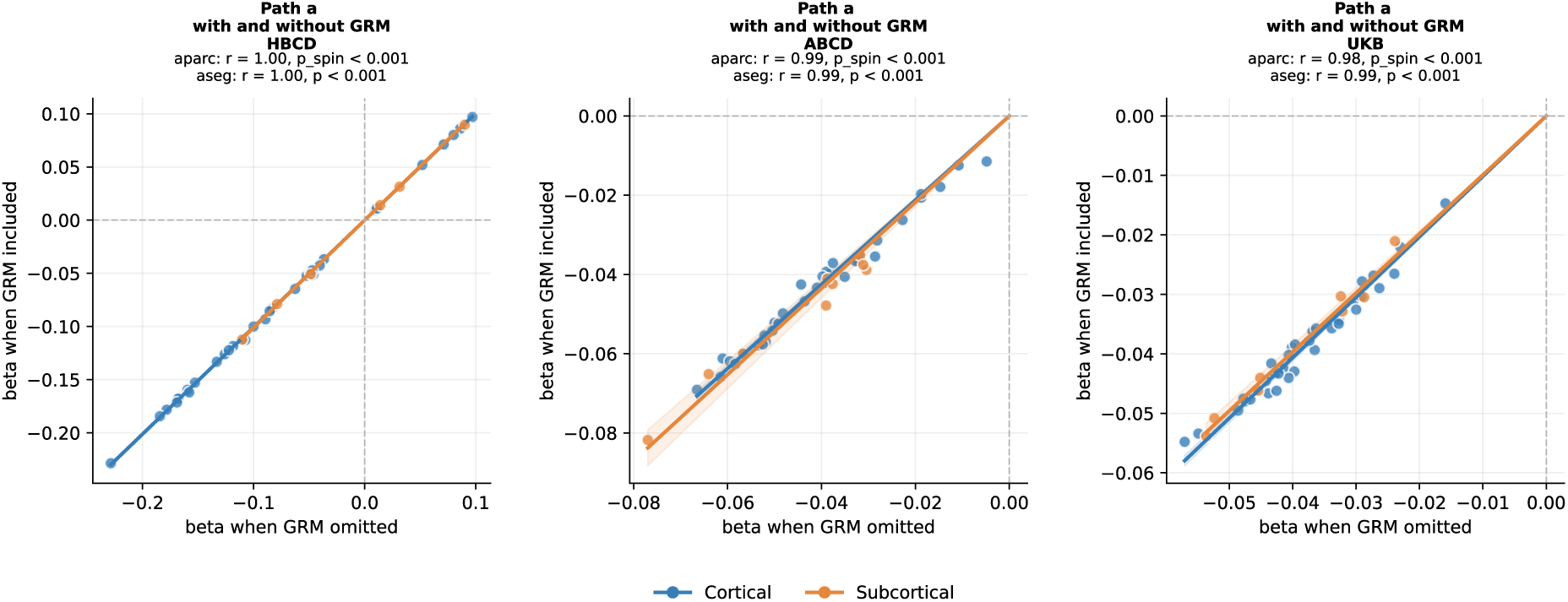
Concordance between primary and kinship-adjusted deprivation effects on brain volume.

